# Violence, firearms, and the coronavirus pandemic: Findings from the 2020 California Safety and Wellbeing Survey

**DOI:** 10.1101/2020.10.03.20206367

**Authors:** Nicole Kravitz-Wirtz, Amanda Aubel, Julia Schleimer, Rocco Pallin, Garen Wintemute

## Abstract

**IMPORTANCE:** Violence is a significant public health problem that has become entwined with the coronavirus pandemic. Conditions that contribute to violence—poverty, unemployment, lack of available resources, isolation, hopelessness, and loss—have intensified and are further compounded by the recent surge in firearm sales, which is itself a risk factor for firearm-related harm.

**OBJECTIVE:** To describe individuals’ worry about violence for themselves and others in the context of the pandemic, pandemic-related unfair treatment, as well as the prevalence of and reasons for firearm acquisition and changes in firearm storage practices due to the pandemic. DESIGN, SETTING, AND PARTICIPANTS: This cross-sectional study used data from the California Safety and Wellbeing Survey, a statewide Internet survey of 2,870 California adults (18 years of age and older) conducted from July 14 to July 27, 2020. Responses were weighted to be representative of the state population of adults.

**MAIN OUTCOMES AND MEASURES:** Survey topics for this study included: changes in worry about violence happening to oneself, by type of violence and location, before and during the pandemic; concern someone else might physically hurt another person or themselves on purpose due to a pandemic-related loss; experiences of unfair treatment related to the pandemic; firearm and ammunition acquisition and changes in firearm storage practices due to the pandemic; and participation in civic and political activities “in response to gun violence” during the pandemic.

**RESULTS:** Worry about violence significantly increased during the pandemic for all violence types except mass shootings. More than 1 in 10 respondents were concerned that someone they know might intentionally harm another person (12.2%) or themselves (13.1%). Of those concerned about self-harm for someone else, 7.5% said it was because the person had suffered a pandemic-related loss. An estimated 110,000 individuals acquired a firearm in response to the pandemic (2.4% of current firearm owners), including 47,000 new owners. Approximately 55,000 individuals (1.2% of owners) who currently store at least one firearm loaded and not locked up said they had adopted this unsecure storage practice in response to the pandemic.

**CONCLUSIONS AND RELEVANCE:** Given the impulsive nature of many types of violence, short-term crisis interventions, such as options for temporary firearm storage outside the home, extreme risk protection orders, and efforts involving community-based violence intervention workers, may be critical for reducing violence-related harm now and following other societal shocks.

**Key Points:** *QUESTIONS:* Has the coronavirus pandemic modified (1) individuals’ worry about violence for themselves or others, (2) the prevalence of and reasons for firearm and ammunition acquisition, and (3) firearm storage practices?

*FINDINGS:* In this cross-sectional, population-representative survey of 2,870 adults in California, worry about multiple types of violence for oneself increased during the pandemic; pandemic-related loss contributed to concern that someone else might physically harm themselves on purpose; an estimated 110,000 people acquired firearms due to the pandemic (2.4% of firearm owners in the state), including approximately 47,000 new owners; and 6.7% of owners who currently store firearms loaded and not locked up adopted this unsecure storage practice in response to the pandemic.

*MEANING:* Violence is a significant public health problem that touches the lives of far more people than is typically recognized. The coronavirus pandemic and efforts to lessen its spread have compounded this burden.

## INTRODUCTION

In the United States (US), in 2018, there were nearly 68,000 violence-related deaths.^1^ An additional 3.3 million people reported having been victims of nonfatal violent crime.^2^ Most deaths (57%) and nearly 471,000 nonfatal violent victimizations involved a firearm.^1,2^ Black, Indigenous, and people of color endure a disproportionate share of this burden.^3-5^ It is reasonable to expect that the emergence and progression of the coronavirus pandemic—with nearly 7.2 million confirmed cases and over 205,000 deaths nationally as of September 30, 2020^6^—combined with the social, psychological, and economic fallout associated with efforts to lessen its spread, are intensifying violence-related harms and inequities therein.

The pandemic has exacerbated longstanding injustices rooted in systemic racism and other oppressive systems of power that contribute to the underlying conditions (e.g., poverty, unemployment, lack of available resources) that elevate risk for, and compound the consequences of, community violence.^7-10^ Recent, largely peaceful,^11^ protests decrying structural inequities, which simultaneously allow police violence and the uneven burden of disease to persist, have been met, at times, by law enforcement use of crowd-control weapons^12^ and heavily armed white supremacist and far-right vigilantes.^13^ Pandemic-induced social isolation, hopelessness, and loss, particularly for people with existing mental health problems such as depression, may result in thoughts of suicide. Violence in the home may increase in frequency and severity as household members, including intimate partners, children, and vulnerable elders, spend more time at home together under high-stress conditions. Having a firearm readily available in these situations creates additional risk.^14-17^

While most major news sources reported initial decreases in violent crime, as measured by local police calls for service, following pandemic-related lockdowns and stay-at-home orders, the latest indications are that more serious acts of violence, particularly those involving firearms, have remained the same or increased.^18^ In addition to a marked rise in shootings in several big cities across the country,^19^ the pandemic appears to have fueled a surge in firearm background checks, an established proxy for firearm sales. Previous spikes in purchasing, such as those following mass shootings and political elections, have been associated with increased firearm violence.^20^ Similarly, recent research suggests an excess of 2.1 million firearm purchases during the first three months of the pandemic, corresponding with an additional 216 to 1,335 fatal and nonfatal firearm-related injuries nationwide.^21^

However, the lack of self-report data capturing individuals’ lived experiences of violence in the context of the pandemic has limited our understanding of the intersection of these coinciding public health problems. One of the only studies to survey individuals who purchased a firearm due to the pandemic relied on a non-representative sample of respondents who did not reflect the socio-demographic profile of most firearm owners.^22^ The current study provides what is to our knowledge the first population-representative estimates of individuals’ worry about violence for themselves, before and during the pandemic; concern someone they know might harm themselves or others due to a pandemic-related loss; experiences of unfair treatment related to the pandemic; firearm and ammunition purchasing and changes in firearm storage practices due to the pandemic; and civic or political activities undertaken “in response to gun violence” during the pandemic.

## METHODS

Data for this cross-sectional survey study come from the 2020 California Safety and Wellbeing Survey, a statewide survey designed by the University of California Firearm Violence Research Center and the Violence Prevention Research Program, both at the University of California Davis, and administered online from July 14 to July 27, 2020 by Ipsos Public Affairs, LLC (Ipsos). The survey was approved by the University of California Davis Institutional Review Board.

Respondents were drawn from KnowledgePanel, an online survey research panel with approximately 60,000 members who are randomly recruited on an ongoing basis through probability-based sampling. Panel members who were aged 18 years and older and residents of California, except those currently serving in the US Armed Forces, were eligible to participate.

Invitations were sent by e-mail; automatic reminders were e-mailed to non-responders three days later. Of 5,018 panel members invited to participate, 2,870 completed the survey, yielding a 57% completion rate. The median survey completion time was 26 minutes.

A final survey weight variable provided by Ipsos adjusts for the initial probability of selection into KnowledgePanel and for survey-specific non-response and over- or under-coverage using post-stratification raking ratio adjustments based on cross-classifications of age, gender, race-ethnicity, education, household income, language proficiency, and California region. The weighted sample is representative of the noninstitutionalized adult population of California as reflected in the 2018 American Community Survey.

Survey questions for this study covered five broad domains: (1) worry about violence happening to oneself, by type of violence (homicide, suicide, mass shooting, assault, robbery, police violence, accidental shooting, and stray bullet shooting) and incident location, before and during the pandemic; (2) concern that someone else might physically harm another person or themselves in response to a pandemic-related loss; (3) experiences of unfair treatment related to the pandemic; (4) firearm and ammunition acquisition and firearm storage practices (among current firearm owners) in response to the pandemic; and (5) participation in civic and political activities “in response to gun violence” during the pandemic. Detailed survey items and response options are in **Appendix 1**. Sociodemographic information was collected as part of ongoing panel membership and merged with survey responses.

### Statistical Analysis

To generate statewide prevalence estimates, we calculated weighted percentages and 95% confidence intervals (CI) for each measure or cross-tabulation of measures using the survey and weighting commands in Stata v15.1 (StataCorp LP, College Station, TX). To evaluate differences in responses during compared with before the pandemic, overall and by respondent characteristics, we fit weighted repeated measures multinomial logistic regression models, including interactions between respondent characteristics and an indicator for the period of reference. The margins command was used to generate prevalence differences and 95% CI.

## RESULTS

Approximately two in five respondents (39.8%: 95% CI: 37.1-42.6) reported that they personally know someone who had tested positive for coronavirus, and 1.3% (95% CI: 0.9-2.0) reported that they themselves had tested positive, while 4.2% (95% CI: 3.2-5.3) reported that they had been sick with coronavirus but had not been tested. Additional sociodemographic and firearm ownership-related characteristics of respondents are in **Appendix 2**.

### Worry about violence

The percentage of respondents who reported that they were somewhat or very worried about violence happening to them significantly increased during the pandemic for all violence types except mass shootings, ranging from a 2.8 percentage point increase for robbery (from 65.5% to 68.2%; p<0.01) to 5.4 percentage points each for police violence (from 45.3% to 50.6%; p<0.001) and unintentional shootings (from 42.7% to 48.0%; p<0.001) and 5.6 percentage points for stray bullet shootings (from 44.5% to 50.0%; p<0.001) (**Table 1**). In contrast, worry about mass shootings declined 4.6 percentage points (from 59.9% to 55.3%; p<0.001) during the pandemic.

**Table 1.** Prevalence of worry about violence, before and during the coronavirus pandemic, by violence type and location, 2020 California Safety and Wellbeing Survey, N=2870

|  | Before the pandemic |  | During the pandemic |  | During - Before |  |
| --- | --- | --- | --- | --- | --- | --- |
|  | % (95% CI) |  | % (95% CI) |  | Difference (p-value) |  |
|  | Not worried | Somewhat/very worried | Not worried | Somewhat/very worried | Not worried | Somewhat/very worried |
| Type |  |  |  |  |  |  |
| Homicide | 54.7 (51.9-57.5) | 44.0 (41.2-46.9) | 51.2 (48.4-54.0) | 47.6 (44.8-50.4) | 3.5 (0.001) | 3.6 (0.001) |
| Suicide | 74.6 (71.9-77.1) | 24.5 (22.0-27.2) | 71.0 (68.3-73.7) | 27.7 (25.1-30.5) | 3.5 (0.001) | 3.2 (0.002) |
| Mass shooting | 39.1 (36.5-41.7) | 59.9 (57.3-62.6) | 43.2 (40.5-46.0) | 55.3 (52.6-58.1) | 4.2 (0.000) | 4.6 (0.000) |
| Assault | 39.9 (37.3-42.6) | 59.1 (56.4-61.8) | 36.5 (33.9-39.2) | 62.4 (59.7-65.1) | 3.4 (0.001) | 3.3 (0.002) |
| Robbery | 33.5 (31.0-36.2) | 65.5 (62.8-68.0) | 30.8 (28.4-33.4) | 68.2 (65.6-70.7) | 2.7 (0.008) | 2.8 (0.008) |
| Police violence | 53.7 (50.8-56.5) | 45.3 (42.5-48.1) | 48.1 (45.4-50.9) | 50.6 (47.8-53.4) | 5.5 (0.000) | 5.4 (0.000) |
| Unintentional shooting | 56.2 (53.3-59.0) | 42.7 (39.9-45.5) | 51.0 (48.2-53.8) | 48.0 (45.3-50.9) | 5.2 (0.000) | 5.4 (0.000) |
| Stray bullet shooting | 54.7 (51.9-57.5) | 44.5 (41.7-47.3) | 48.8 (46.0-51.6) | 50.0 (47.3-52.8) | 5.9 (0.000) | 5.6 (0.000) |
| Location |  |  |  |  |  |  |
| Home | 70.6 (67.9-73.1) | 27.9 (25.4-30.5) | 69.4 (66.7-72.0) | 29.1 (26.5-31.7) | 1.2 (0.294) | 1.2 (0.291) |
| Neighborhood | 50.0 (47.3-52.8) | 48.8 (46.0-51.6) | 46.5 (43.7-49.2) | 52.2 (49.4-55.0) | 3.6 (0.002) | 3.4 (0.003) |
| Somewhere else | 27.2 (24.8-29.7) | 71.3 (68.8-73.7) | 26.7 (24.3-29.1) | 71.7 (69.2-74.1) | 0.1 (0.654) | 0.0 (0.710) |

The percentage of respondents who reported being somewhat or very worried about violence happening to them in their neighborhood significantly increased during the pandemic, from 48.4% to 52.2% (p<0.01); there was no change in worry about violence in the home or somewhere else (**Table 1**). Among those who expressed worry about violence in their home, neighborhood, or somewhere else, before and/or during the pandemic, similar percentages reported that their worry was at least in part due to their spouse or intimate partner (9.7% [95% CI: 7.9-11.9] before and 9.0% [95% CI: 7.3-11.1] during) (**Appendix 3**). Of those, most were somewhat or very willing to ask for help from a domestic violence hotline (75.4%; 95% CI: 65.5-83.3), family member or friend (86.8%; 95% CI: 77.1-92.8), and law enforcement (89.5%; 95% CI: 80.5-94.6) (**Appendices 4-6**).

The share of respondents who reported worry about violence by sociodemographic characteristics and firearm ownership status is in **Appendices 3-17**.

### Concern about violence for others

Of the 12.1% (95% CI: 10.4-14.1) of respondents who reported concern that someone they know might physically hurt another person on purpose, 1.8% (95% CI: 0.7-4.4) reported that their concern was at least in part because the person had suffered a major loss (e.g., loss of someone they cared about, a job, or housing) that was related to the pandemic (**Table 2**). Likewise, but more pronounced, of the 13.3% (95% CI: 11.5-15.3) of respondents who reported concern that someone they know might physically hurt themselves on purpose, 7.5% (95% CI: 4.5-12.2) reported that their concern was at least in part because the person had suffered a pandemic-related loss.

**Table 2.** Prevalence of respondents concerned that someone they know might physically hurt another person or themselves on purpose, reasons for concern, and firearm access, California Safety and Wellbeing Survey, N=2870

|  |  |  |
| --- | --- | --- |
|  | Another person | Themselves |
|  | % (95% CI) | % (95% CI) |
| Total | 12.1 (10.4-14.1) | 13.3 (11.5-15.3) |
| Reasons for concern |  |  |
| Pandemic-related loss | 1.8 (0.7-4.4) | 7.5 (4.5-12.2) |
| Firearm access |  |  |
| Yes | 0 | 6.0 (1.2-25.8) |
| No | 10.2 (1.8-41.1) | 36.6 (17.2-61.6) |
| Don't know | 89.8 (58.9-98.2) | 57.4 (32.9-78.7) |
| Other non-pandemic reasons only | 98.2 (95.6-99.3) | 92.5 (87.8-95.5) |

Among respondents whose concerns were due to a pandemic-related loss, most said they did not know whether the other person had access to a firearm (89.8% [95% CI: 58.9-98.2] for other-directed harm and 57.4% [95% CI: 32.9-78.7] for self-harm); 6.0% (95% CI:1.2-25.8) of respondents who were concerned that someone they know might harm themselves due to a pandemic-related loss said the person had access to a firearm (**Table 2**).

### Experiences of unfair treatment

More than two-thirds of respondents (69.2%; 95% CI: 66.6-71.7) reported that they had experienced at least one form of unfair treatment in the past 12 months (**Table 3**). Of those, 7.4% (95% CI: 5.6-9.6) said the unfair treatment was related to the pandemic. Asian respondents most often reported pandemic-related unfair treatment: 17.2% (95% CI: 10.2-27.5) of Asian respondents who experienced unfair treatment said it was related to the pandemic, compared with 10.7% (95% CI: 2.2-39.5) of those who identified as multiracial or other race, 7.5% (95% CI: 2.4-20.5) of Black respondents, 7.4% (95% CI: 5.1-10.7) of white respondents, and 3.0% (95% CI: 1.5-5.9) of Latinx respondents.

**Table 3.** Prevalence of unfair treatment in the past 12 months, overall and due to the pandemic, by race, California Safety and Wellbeing Survey, N=2870

|  |  |  |
| --- | --- | --- |
|  | Experienced unfair treatment | Coronavirus-related unfair treatment |
|  | % (95% CI) | % (95% CI) |
| Total | 69.2 (66.6-71.7) | 7.4 (5.6-9.6) |
| Asian | 66.9 (58.4-74.4) | 17.2 (10.2-27.5) |
| Black | 75.9 (63.8-85.0) | 7.5 (2.4-20.5) |
| Latinx | 69.1 (64.3-73.5) | 3.0 (1.5-5.9) |
| Multiracial/other | 69.2 (50.7-83.1) | 10.7 (2.2-39.5) |
| White | 69.1 (65.7-72.3) | 7.4 (5.1-10.7) |

### Firearm acquisition and storage practices

Nearly one in four respondents (23.5%; 95% CI: 21.3-25.9) reported that they or someone else in their household owned firearms; 15.2% (95% CI: 13.4-17.2) of respondents reported that they were a firearm owner (**Appendix 1**). Among owners, 2.4% (95% CI: 1.1-5.0) reported that they had acquired a firearm in response to the pandemic, while 8.5% (95% CI: 5.0-14.0) of owners, including all of those who had acquired a firearm, said that they had purchased ammunition in response to the pandemic (**Table 4**). Among those who had acquired a firearm in response to the pandemic, 43.0% (95% CI: 14.8-76.6) reported that they did not already own a firearm. Extrapolating to the population of adults in California (30.1 million in 2018), we estimate approximately 110,000 Californians acquired firearms in response to the pandemic, including 47,000 new owners.

**Table 4.** Prevalence of firearm and ammunition acquisition in response to the pandemic and related characteristics among firearm owners, 2020 California Safety and Wellbeing Survey, N=529

|  |  |  |
| --- | --- | --- |
|  | Firearms | Ammunition |
|  | % (95% CI) | % (95% CI) |
| Acquired in response to the pandemic | 2.4 (1.1-5.0) | 8.5 (5.0-14.0) |
| Did not already own a firearm | 43.0 (14.8-76.6) |  |
| Reasons for acquisition |  |  |
| Lawlessness | 75.9 (27.6-96.3) | 77.1 (46.6-92.8) |
| Prisoner releases | 56.1 (22.0-85.3) | 49.2 (24.6-74.2) |
| Government going too far | 49.2 (17.7-81.3) | 38.5 (17.8-64.4) |
| Government collapse | 38.0 (12.2-73.0) | 52.9 (27.6-76.7) |
| Gun stores closing | 31.1 (9.7-65.4) | 35.8 (15.3-63.3) |
| Hunting | 10.5 (1.4-49.4) | 7.2 (1.8-24.6) |
| Sport shooting | 10.5 (1.4-49.4) | 8.1 (2.7-22.2) |
| Some other reason | 0 | 4.0 (0.9-15.9) |
| Firearm storage |  |  |
| All guns unloaded and locked up | 62.7 (56.2-68.7) |  |
| Change in response to the pandemic | 0 |  |
| ≥1 gun(s) loaded and not locked up | 18.0 (13.1-24.1) |  |
| Change in response to the pandemic | 6.7 (2.7-15.6) |  |
| Some other way | 18.6 (14.8-23.1) |  |
| Note: Reasons for acquisition are not mutually exclusive |  |  |

The most common reason given for firearm acquisition in response to the pandemic was worry about lawlessness (75.9%; 95% CI: 27.6-96.3), followed by worry about prisoner releases (56.1; 95% CI: 22.0-85.3), the government going too far (49.2%; 95% CI: 17.7-81.3), government collapse (38.0%; 95% CI: 12.2-73.0), and gun stores closing (31.1%; 95% CI: 9.7-65.4) (**Table 4**). Reasons for ammunition purchases in response to the pandemic were similar. Firearm owners (vs non-owners) and those who had acquired a firearm in response to the pandemic (vs non-owners and owners without a pandemic-related acquisition) also had the largest percentage increases in their level of worry about multiple types of violence during (vs before) the pandemic (**Appendices 7-17**).

Among firearm owners, 62.7% (95% CI: 56.2-68.7) reported that they currently store all of their firearms in the most secure way (i.e., unloaded and locked up), 18.0% (95% CI: 13.1-24.1) store at least one firearm in the least secure way (i.e., loaded and not locked up), and the remainder store their firearms in some other way (18.6%; 95% CI: 14.8-23.1) (e.g., unloaded but not locked up) (**Table 4**). Of owners who currently store at least one firearm in the least secure way, 6.7% (95% CI: 2.7-15.6) reported that this reflected a change in storage practice due to the pandemic. Of those, approximately half (53.0%; 95% CI: 17.2-86.0) lived in households with children or teens.

### Civic and political activity

More than half (52.4%; 95% CI: 50.0-55.2) of respondents reported that they had done one or more civic or political activities “in response to gun violence” in the past 12 months (**Table 5**). Of those, the plurality reported that they had read about a political candidate’s position (40.7%; 95% CI: 38.1-43.4). Nearly one in four respondents (22.2%; 95% CI: 19.4-25.3) who reported having done something in the past 12 months (11.6% [95% CI: 10.1-13.4] of adults in the state) said that they had also done something in the past 2 months, during the pandemic and protests for racial justice and police accountability.

**Table 5.**
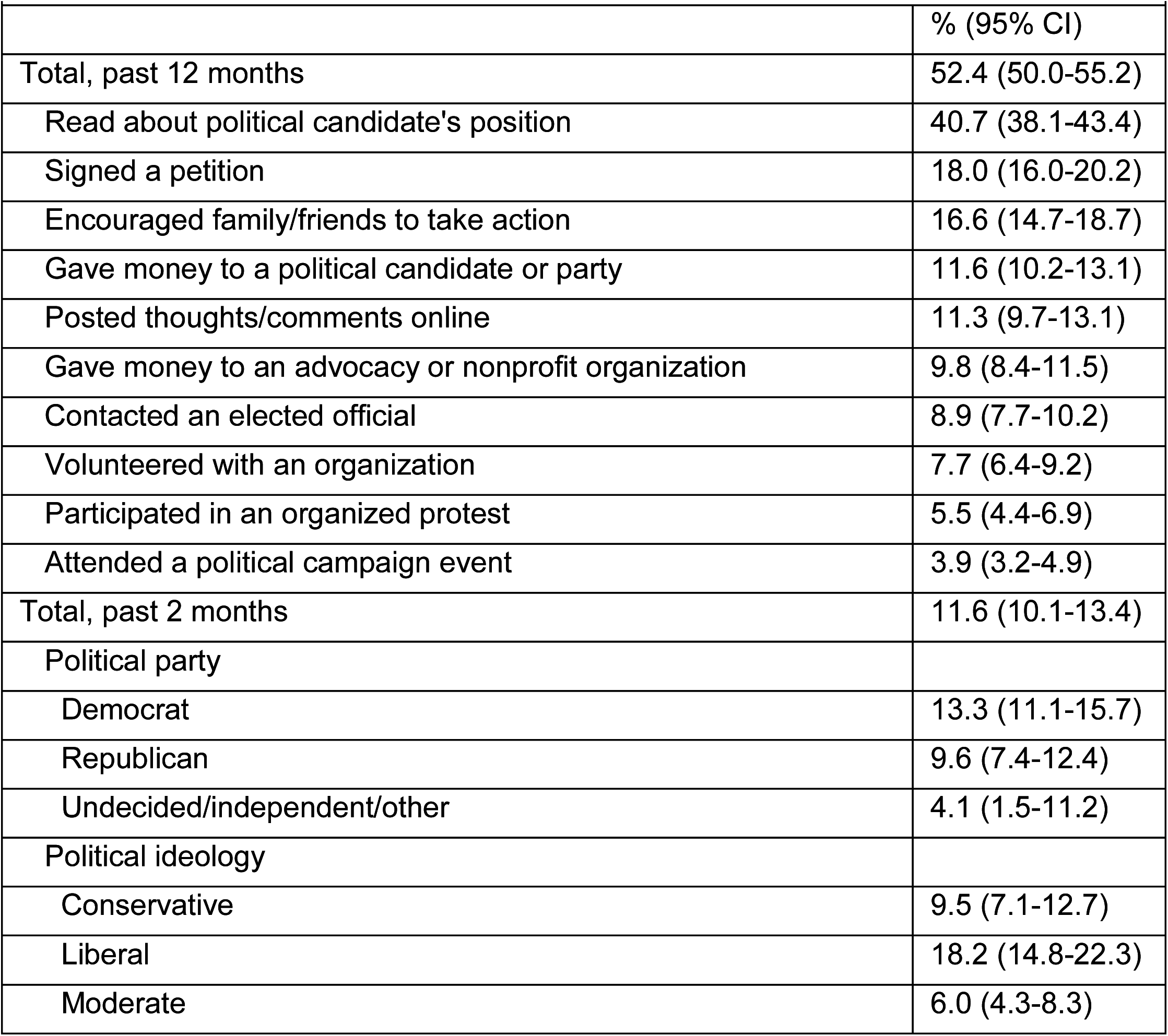
Prevalence of participation in civic or political activities in response to gun violence in America, California Safety and Wellbeing Survey, N=2870

## DISCUSSION

Violence is a significant public health problem which touches the lives of far more people than is typically recognized; violence affects people not only through direct involvement, but also through indirect and vicarious experiences that ripple across individuals, families, and entire communities. Our findings from this first-of-its-kind population-representative survey of California adults add support to a growing body of research suggesting that the coronavirus pandemic and efforts to lessen its spread have compounded the burden of violence-related harms.

Our respondents expressed increased levels of worry about violence during compared with before the pandemic, ranging from 2.8 to 5.6 percentage point increases in the estimated statewide prevalence of adults who reported that they were somewhat or very worried about multiple types of interpersonal violence (i.e., robbery, assault, homicide, police violence), suicide, and unintentional firearm injury happening to them. Pandemic-related experiences of unfair treatment were also reported and disproportionately common among Asian Americans. In addition, more than one in ten respondents were concerned that someone they know might physically harm another person (12.1%) or themselves (13.3%) on purpose. The role of the pandemic was particularly pronounced for those concerned about self-harm for someone else: 7.5% said their concern was at least in part because the person had suffered a pandemic-related loss. In 6.0% of these cases, the person had known access to a firearm.

Emerging research also indicates a surge in firearm background checks in the months coinciding with the pandemic,^21^ though we are among the first to estimate the prevalence of and motivations for ammunition and firearm acquisition in direct response to the pandemic. We found that approximately 1 in 12 (8.5%) firearm owners in California purchased ammunition in response to the pandemic, including roughly 110,000 individuals who also acquired firearms (2.4% of owners in the state). Of those, an estimated 47,000 were new owners, who may have little past experience or training with firearms. Previous spikes in firearm purchasing have been associated with increased firearm violence,^20^ and recent evidence suggests a similar relationship exists during the pandemic.^21^

Consistent with the most common reasons for firearm ownership generally,^23,24^ respondents who acquired firearms in response to the pandemic usually did so for self-protection: three-quarters (75.9%) indicated worry about lawlessness and more than half (56.1%) endorsed worry about prisoner releases. Although the perceived need for self-protection continues to motivate firearm ownership amid the pandemic, an extensive body of evidence suggests instead that the presence of a firearm in the home elevates risk for firearm-related harm, particularly unintentional shootings (often involving children), female homicide victimization, and completed suicide.^14-17^ More concerning, perhaps, is that people who own firearms primarily for protection are more likely to store firearms in the home loaded and/or not locked up,^25^ an independent risk factor for firearm injury and death. Our findings suggest the pandemic may contribute to this risk: an estimated 55,000 people (1.2% of owners in the state) who currently store at least one firearm loaded and not locked up reported adopting this unsecure storage practice in response to the pandemic.

Taken together, our findings add support to long-term public health-oriented prevention and intervention strategies designed to address the enduring psychological trauma associated with exposure to and worry about violence, as well as the intermediary (e.g., firearm ownership and storage) and upstream (e.g., socioeconomics, education, the environment) determinants of violence risk. More immediately, given the impulsive nature of many types of violence, and the multiple acute disruptions associated with the pandemic, short-term crisis interventions, such as options for temporary firearm storage outside the home, extreme risk protection orders, and efforts involving community-based violence intervention workers, may be particularly critical for reducing the burden of violence. As underscored in our results, many respondents (52.4%) have already taken action in response to this prevailing public health problem.

### Limitations

This study has some limitations. First, we rely on self-report data, which is subject to social desirability, non-response, and recall biases. However, several administrative data sources provide an opportunity to broadly assess the validity of our estimates. The Johns Hopkins Coronavirus Resource Center documented 346,000 to 458,000 confirmed cases of coronavirus in California at the time our survey was in the field, close to our 391,000 estimate after extrapolating to the population of adults in the state. Similarly, National Instant Criminal Background Check System (NICS) data show roughly 557,000 people underwent a firearm background check in California from March through June 2020, compared with 465,000 during the same period in 2019. This amounts to a year-over-year increase of 92,000, close to our 110,000 estimate of pandemic-related firearm acquisitions.

Second, given state-level differences in infection rates and in efforts to lessen the spread and impacts of coronavirus, as well as California’s relatively low rates of firearm ownership and more comprehensive firearm regulations, our findings might not be generalizable to other states. However, coronavirus is a near ubiquitous exposure across the US and nationally-representative studies have similarly found deleterious impacts of the pandemic on psychological health,^26^ as well as nationwide pandemic-related spikes in firearm purchasing.^21^

Third, we use a retrospective pre-post approach to compare responses before and during the pandemic, which may inaccurately reflect the impacts of the pandemic if respondents’ knowledge or experiences associated with the pandemic led them to interpret questions in a qualitatively different manner. However, some research suggests that when individuals are asked to respond to questions about a particular subject after they have some basic knowledge of or experience with the subject itself, they are better able to accurately reflect on the degree of change.^27^

## CONCLUSIONS

The coronavirus pandemic has exacerbated persistent structural, economic, and social inequities in the conditions that contribute to violence and its consequences. Findings from this study assessing the near-term effects of the pandemic on individual perceptions, motivations, and behaviors related to violence and firearm ownership can inform prevention and intervention efforts now and following other societal shocks, as well as lay the groundwork for more comprehensive research and prevention efforts in the future.

## Data Availability

Data is not publicly available.

## REFERENCES

1. CDC. Fatal injury reports. Web-based Injury Statistics Query and Analysis System (WISQARS). 2018. Accessed 08/31/2020.

2. National Crime Victimization Survey (NCVS), 2018. Bureau of Justice Statistics.

3. Kalesan B, Vyliparambil MA, Zuo Y, et al. Cross-sectional study of loss of life expectancy at different ages related to firearm deaths among black and white Americans. BMJ Evidence-Based Medicine. 2019;24(2):55.

4. Edwards F, Lee H, Esposito M. Risk of being killed by police use of force in the United States by age, race–ethnicity, and sex. Proceedings of the National Academy of Sciences. 2019;116(34):16793.

5. Edwards F, Esposito MH, Lee H. Risk of police-involved death by race/ethnicity and place, United States, 2012–2018. American journal of public health. 2018;108(9):1241–1248.

6. Johns Hopkins Coronavirus Resource Center. https://coronavirus.jhu.edu/. Accessed 9/30/2020.

7. Jacoby SF, Dong B, Beard JH, Wiebe DJ, Morrison CN. The enduring impact of historical and structural racism on urban violence in Philadelphia. Social Science & Medicine. 2018;199:87–95.

8. Rowhani-Rahbar A, Quistberg DA, Morgan ER, Hajat A, Rivara FP. Income inequality and firearm homicide in the US: a county-level cohort study. Injury prevention. 2019;25(Suppl 1):i25–i30.

9. Bryan A, Diuguid-Gerber J, Davis NJ, Chokshi DA, Galea S. Moving From The Five Whys To Five Hows: Addressing Racial Inequities In COVID-19 Infection And Death. Health Affairs Blog.

10. Pollack CE, Leifheit KM, Linton SL. When Storms Collide: Evictions, COVID-19, And Health Equity. Health Affairs Blog.

11. The Armed Conflict Location & Event Data Project (ACLED). Demonstrations & Political Violence in America: New Data for Summer 2020. https://acleddata.com/2020/09/03/demonstrations-political-violence-in-america-new-data-for-summer-2020/.

12. Physicians for Human Rights. Shot in the Head. September 14, 2020.

13. German M. Hidden in Plain Sight: Racism, White Supremacy, and Far-Right Militancy in Law Enforcement. Brennan Center for Justice;2020.

14. Studdert DM, Zhang Y, Swanson SA, et al. Handgun Ownership and Suicide in California. New England Journal of Medicine. 2020;382(23):2220–2229.

15. Anglemyer A, Horvath T, Rutherford G. The accessibility of firearms and risk for suicide and homicide victimization among household members: a systematic review and meta-analysis. Annals of internal medicine. 2014;160(2):101–110.

16. Hemenway D. Risks and Benefits of a Gun in the Home. American Journal of Lifestyle Medicine. 2011;5(6):502–511.

17. Miller M, Azrael D, Hemenway D. Firearm availability and unintentional firearm deaths, suicide, and homicide among 5–14 year olds. Journal of Trauma and Acute Care Surgery. 2002;52(2):267–275.

18. Ashby MP. Initial evidence on the relationship between the coronavirus pandemic and crime in the United States. Crime Science. 2020;9:1–16.

19. Sutherland M, McKenney M, Elkbuli A. Gun violence during COVID-19 pandemic: Paradoxical trends in New York City, Chicago, Los Angeles and Baltimore. The American journal of emergency medicine. 2020:S0735-6757(0720)30344-30342.

20. Laqueur HS, Kagawa RM, McCort CD, Pallin R, Wintemute G. The impact of spikes in handgun acquisitions on firearm-related harms. Injury epidemiology. 2019;6(1):1–6.

21. Schleimer JP, McCort CD, Pear VA, et al. Firearm Purchasing and Firearm Violence in the First Months of the Coronavirus Pandemic in the United States. medRxiv. 2020:2020.2007.2002.20145508.

22. Lyons RA, Finch CF, McClure R, van Beeck E, Macey S. The injury List Of All Deficits (LOAD) Framework – conceptualising the full range of deficits and adverse outcomes following injury and violence. International Journal of Injury Control and Safety Promotion. 2010;17(3):145–159.

23. Kravitz-Wirtz N, Pallin R, Miller M, Azrael D, Wintemute GJ. Firearm ownership and acquisition in California: findings from the 2018 California Safety and Well-being Survey. Injury Prevention. 2019:injuryprev-2019-043372.

24. Azrael D, Hepburn L, Hemenway D, Miller M. The Stock and Flow of U.S. Firearms: Results from the 2015 National Firearms Survey. RSF. 2017;3(5):38–57.

25. Schleimer JP, Kravitz-Wirtz N, Pallin R, Charbonneau AK, Buggs SA, Wintemute GJ. Firearm ownership in California: A latent class analysis. Injury Prevention. 2019:injuryprev-2019-043412.

26. Ettman CK, Abdalla SM, Cohen GH, Sampson L, Vivier PM, Galea S. Prevalence of Depression Symptoms in US Adults Before and During the COVID-19 Pandemic. JAMA Network Open. 2020;3(9):e2019686–e2019686.

27. Rockwell SK, Kohn H. Post-then-pre evaluation. Journal of Extension. 1989;27(2):19–21.

