## Supplemental Materials for "Violence, firearms, and the coronavirus pandemic: Findings from the 2020 California Safety and Wellbeing Survey"

Q3. **[Accordion Grid]**
In general, **before** the coronavirus epidemic, how worried were you about each of the following violent events happening to **you**?

*Scripter: Randomize order of statements. Record order in which statements were shown.*

*Statements:*
1. Homicide
2. Suicide
3. Mass shooting
4. Assault
5. Robbery
6. Police violence
7. Accidental shooting
8. Getting hit by a stray bullet

*Responses:*
1. Not worried
2. Somewhat worried
3. Very worried 17

Q4. **[Accordion Grid]**
These days, **during** the coronavirus epidemic, how worried are you about each of the following violent events happening to **you**?

*Scripter: Same order of items as Q3*

*Statements:*
1. Homicide
2. Suicide
3. Mass shooting
4. Assault
5. Robbery
6. Police violence
7. Accidental shooting
8. Getting hit by a stray bullet

*Responses:*
1. Not worried
2. Somewhat worried
3. Very worried

Q7. **[Grid, SP Across]**
In general, **before** the coronavirus epidemic, how worried were you about a violent event happening to **you** in each of the following places?

*Statements per row:*
1. In my home
2. Not in my home but in my neighborhood
3. Somewhere else

*Answers in columns:*
1. Not worried
2. Somewhat worried
3. Very worried

Q8. **[S]**
You said you were somewhat or very worried about a violent event happening to **you**. Was this worry at least in part because of your spouse or intimate partner?

1. Yes
2. No 18

Q9. **[Grid, SP Across]**
These days, **during** the coronavirus epidemic, how worried are you about a violent event happening to **you** in each of the following places?

*Statements per row:*
1. In my home
2. Not in my home but in my neighborhood
3. Somewhere else

*Answers in columns:*
1. Not worried
2. Somewhat worried
3. Very worried

Q10. **[S]**
You said you are somewhat or very worried about a violent event happening to **you**. Is this worry at least in part because of your spouse or intimate partner?

1. Yes
2. No

Q12. **[Accordion Grid]**
How willing are you to ask for help from the following people?

*Statements:*
1. A domestic violence hotline
2. A family member or trusted friend
3. Law enforcement

*Responses:*
1. Not at all willing
2. Somewhat willing
3. Very willing

Q35. **[S]** [PROMPT ONCE IF NO RESPONSE GIVEN]
Are you concerned that anyone you know might physically hurt **another person** on purpose? Consider only people you know personally, not people you've only heard about from others or seen in the media.

1. Yes
2. No

Q47. **[M]**
Do any of these things make you concerned this person might physically hurt another person on purpose? Select all that apply.

1. They have suffered a major loss, such as loss of someone they cared about, loss of a job, or loss of housing
2. They have hurt another person before
3. They have hurt themselves before
4. They have trouble with alcohol or drug use
6. They have posted about violence or made threats to hurt another person on social media
7. They have talked about violence or made threats to hurt another person
10. They have damaged property
11. Other, please specify: [Text box]
12. None of the above [E]

Q48. **[S]**
You said they have suffered a major loss. Was this loss related to the coronavirus epidemic or something else?

1. The coronavirus epidemic
2. Something else
3. Both
4. Don’t know

Q52. **[S]**
Does this person have access to a gun?

1. Yes
2. No
3. Don't know

Q63. **[S]** [PROMPT ONCE IF NO RESPONSE GIVEN]
Are you concerned that anyone you know might physically hurt **themselves** on purpose? Consider only people you know personally, not people you've only heard about from others or seen in the media.

1. Yes
2. No

Q75. **[M]**
Do any of these [IF Q72 = 1 AND/OR Q74 = 1, INSERT: other] things make you concerned this person might physically hurt themselves on purpose? Select all that apply.

1. They have suffered a major loss, such as loss of someone they cared about, loss of a job, or loss of housing
2. They have hurt another person before
3. They have hurt themselves before
4. They have trouble with alcohol use or drug use
6. They have posted about violence or made threats to hurt themselves on social media
7. They have talked about violence or made threats to hurt themselves
10. They have damaged property
11. Other, please specify:[Text box] [A]
12. None of the above [A,E]

Q76. **[S]**
You said they have suffered a major loss. Was this loss related to the coronavirus epidemic, or something else?

1. The coronavirus epidemic
2. Something else
3. Both
4. Don't know

Q78. **[S]**
Does this person have access to a gun?

1. Yes
2. No
3. Don't know

Q129. **[Accordion Grid]**
In the past 12 months, how often have any of the following things happened to you?

*Statements per row:*
1. You were treated with less respect than other people
2. You were treated unfairly at restaurants or stores or other public places
3. People criticized your accent or the way you speak
4. People acted as if they think you are not smart
5. People acted as if they are afraid of you
6. People acted as if they think you are dishonest
7. People acted as if they think they are better than you are
8. You were threatened or harassed
9. You were physically assaulted

*Answers in columns:*
1. Never
2. Rarely
3. Sometimes
4. Often
5. Don't know

Q132. **[S]** [PROMPT ONCE IF NO RESPONSE GIVEN]
Was any of the unfair treatment related to the coronavirus epidemic?

1. Yes
2. No
3. Don't know

Q110. **[S]** [PROMPT ONCE IF NO RESPONSE GIVEN]
Have you acquired any guns in response to the coronavirus epidemic?

1. Yes
2. No

Q111. **[M]**
You said you acquired a gun in response to the coronavirus epidemic. Did you get this gun for any of the specific reasons below? Select all that apply.

1. I'm worried about lawlessness
2. I'm worried about government collapse
3. I'm worried about government going too far
4. I'm worried about prisoner releases
5. I'm worried about getting enough food and need to hunt
6. I have more time on my hands for sport shooting
7. I'm worried about gun stores being closed
8. Other, please specify ___________ [A] [SCRIPTER: PROMPT ONCE IF SELECTED AND NO RESPONSE GIVEN]

Q112. **[S]**
You said you acquired a gun in response to the coronavirus epidemic. Did you already personally own a gun at that time?

1. Yes
2. No

Q113. **[S]** [PROMPT ONCE IF NO RESPONSE GIVEN]
Have you bought ammunition in response to the coronavirus epidemic?

1. Yes
2. No 45

Q114. **[M]**
You said you bought ammunition in response to the coronavirus epidemic. Did you buy this ammunition for any of the specific reasons below? Select all that apply.

1. I'm worried about lawlessness
2. I'm worried about government collapse
3. I'm worried about government going too far
4. I'm worried about prisoner releases
5. I'm worried about getting enough food and need to hunt
6. I have more time on my hands for sport shooting
7. I'm worried about gun stores being closed
8. Other, please specify ___________ [A] [SCRIPTER: PROMPT ONCE IF SELECTED AND NO RESPONSE GIVEN]

Q115. **[M]**
How do you currently store your gun(s)? Select all that apply.

1. Unloaded and locked up (for example, with a trigger lock, cable lock, in a lock box or safe)
2. Loaded and not locked up
3. Some other way

Q116. **[S]**
You said you stored gun(s) unloaded and locked up. Have you always stored gun(s) this way?

1. Yes
2. No

Q117. **[S]**
Is this a change because of the coronavirus epidemic?

1. Yes
2. No

Q118. **[S]**
You said you stored gun(s) loaded and not locked up. Have you always stored gun(s) this way?

1. Yes
2. No

Q119. **[S]**
Is this a change because of the coronavirus epidemic?

1. Yes
2. No

eAppendix 2. Respondent characteristics, California Safety and Wellbeing Survey 2020, n=2870

| | n | % [95% CI] |
|---|---|---|
| Age | | |
| 18-29 | 200 | 16.3 [14.0-18.9] |
| 30-44 | 511 | 29.7 [27.0-32.5] |
| 45-59 | 738 | 26.7 [24.4-29.2] |
| 60+ | 1421 | 27.3 [25.3-29.4] |
| Race/ethnicity | | |
| White, Non-Hispanic | 1615 | 41.9 [39.3-44.6] |
| Black, Non-Hispanic | 127 | 5.8 [4.6-7.3] |
| Asian, Non-Hispanic | 208 | 14.4 [12.3-16.8] |
| Other & 2+ races, Non-Hispanic | 71 | 3.2 [2.3-4.5] |
| Hispanic | 849 | 34.7 [32.0-37.4] |
| Gender | | |
| Male | 1506 | 47.7 [45.0-50.5] |
| Female | 1364 | 52.3 [49.5-55.0] |
| Education | | |
| Less than high school | 158 | 15.4 [13.1-18.0] |
| High school | 364 | 20.4 [18.1-23.0] |
| Some college | 938 | 32.7 [30.2-35.2] |
| Bachelor's degree | 775 | 17.5 [15.7-19.4] |
| Advanced degree | 635 | 14.0 [12.4-15.8] |
| Income | | |
| Less than $25,000 | 431 | 15.1 [13.2-17.2] |
| $25,000-$59,999 | 744 | 23.9 [21.7-26.3] |
| $60,000-$99,999 | 757 | 22.6 [20.4-24.9] |
| $100,000 or more | 938 | 38.5 [35.8-41.3] |
| Household gun ownership | | |
| No guns in home | 1989 | 71.4 [68.8-73.8] |
| Gun owner | 529 | 8.3 [6.9-10.0] |
| Non-owner living with owner | 219 | 15.2 [13.4-17.2] |
| Acquired firearms due to the coronavirus pandemic | | |
| Non-gun owner | 2341 | 84.8 [82.8-86.6] |
| Gun owner, no purchase | 519 | 14.8 [13.0-16.8] |
| Gun owner, purchase | 10 | 0.4 [0.2-0.8] |

Columns may not sum to 100% due to non-response

eAppendix 3. Percentage of respondents worried about intimate partner violence happening to them before and during the coronavirus pandemic, among those worried about a violent event happening to them, California Safety and Wellbeing Survey 2020

| | Before the pandemic | | | | | | During the pandemic | | | | | |
|---|---|---|---|---|---|---|---|---|---|---|---|---|
| | No | | Yes | | Total | | No | | Yes | | Total | |
| | n | % [95% CI] | n | % [95% CI] | n | % | n | % [95% CI] | n | % [95% CI] | n | % |
| Total | 1881 | 90 [87.9-91.9] | 179 | 9.7 [7.9-11.8] | 2,069 | 100 | 1884 | 90.7 [88.6-92.4] | 176 | 9 [7.3-11.1] | 2,069 | 100 |
| Education | | | | | | | | | | | | |
| Less than high school | 102 | 78.5 [69.2-85.6] | 29 | 21.5 [14.4-30.8] | 131 | 100 | 100 | 79.4 [70.1-86.4] | 25 | 20 [13.1-29.3] | 126 | 100 |
| High school | 255 | 94.5 [91.3-96.6] | 24 | 5.5 [3.4-8.7] | 279 | 100 | 257 | 94.5 [91.4-96.5] | 26 | 5.5 [3.5-8.6] | 283 | 100 |
| Some college | 628 | 92.2 [89.3-94.4] | 50 | 7 [4.9-9.9] | 684 | 100 | 643 | 93 [90.3-95.1] | 48 | 6.5 [4.5-9.2] | 695 | 100 |
| Bachelor's degree | 481 | 89.6 [84.0-93.3] | 49 | 10.3 [6.6-15.9] | 531 | 100 | 486 | 91.2 [86.4-94.5] | 48 | 8.6 [5.4-13.4] | 536 | 100 |
| Advanced degree | 415 | 92.7 [87.1-96.0] | 27 | 7.1 [3.8-12.7] | 444 | 100 | 398 | 91.8 [85.8-95.4] | 29 | 8 [4.4-14.0] | 429 | 100 |
| Gender | | | | | | | | | | | | |
| Male | 898 | 84.6 [80.7-87.8] | 134 | 15.3 [12.0-19.1] | 1,036 | 100 | 905 | 84.8 [81.0-88.0] | 135 | 15 [11.9-18.8] | 1,045 | 100 |
| Female | 983 | 94.7 [92.2-96.4] | 45 | 4.9 [3.3-7.4] | 1,033 | 100 | 979 | 95.8 [93.7-97.2] | 41 | 3.8 [2.4-5.9] | 1,024 | 100 |
| Race/ethnicity | | | | | | | | | | | | |
| White, Non-Hispanic | 1004 | 93.5 [90.7-95.5] | 58 | 6 [4.0-8.7] | 1,068 | 100 | 1012 | 92.9 [89.8-95.1] | 61 | 6.7 [4.5-9.8] | 1,078 | 100 |
| Black, Non-Hispanic | 90 | 96.6 [92.0-98.6] | 5 | 3.1 [1.2-7.9] | 96 | 100 | 80 | 94.3 [88.1-97.4] | 7 | 5.4 [2.4-11.6] | 88 | 100 |
| Asian, Non-Hispanic | 148 | 87.7 [79.8-92.8] | 16 | 12.3 [7.2-20.2] | 164 | 100 | 151 | 89.4 [82.3-93.9] | 16 | 10.6 [6.1-17.7] | 167 | 100 |
| Other & 2+ races, Non-Hispanic | 42 | 97.2 [90.4-99.2] | 3 | 2.8 [0.8-9.6] | 45 | 100 | 44 | 98.7 [94.4-99.7] | 2 | 1.3 [0.3-5.6] | 46 | 100 |
| Hispanic | 597 | 86.1 [81.8-89.4] | 97 | 13.8 [10.4-18.0] | 696 | 100 | 597 | 87.9 [83.9-90.9] | 90 | 11.8 [8.8-15.8] | 690 | 100 |
| Income | | | | | | | | | | | | |
| Less than $25,000 | 303 | 88 [81.2-92.6] | 39 | 11.7 [7.2-18.5] | 343 | 100 | 297 | 88.2 [81.4-92.8] | 36 | 11.1 [6.7-18.0] | 335 | 100 |
| $25,000-$59,999 | 512 | 89.2 [84.8-92.4] | 54 | 10.8 [7.5-15.2] | 567 | 100 | 527 | 89.9 [85.9-92.9] | 54 | 10 [7.0-14.1] | 582 | 100 |
| $60,000-$99,999 | 484 | 91.7 [87.9-94.4] | 39 | 7.6 [5.0-11.5] | 528 | 100 | 481 | 91.7 [87.7-94.4] | 39 | 7.9 [5.2-11.9] | 524 | 100 |
| $100,000 or more | 582 | 90.5 [86.2-93.6] | 47 | 9.2 [6.2-13.5] | 631 | 100 | 579 | 91.7 [87.7-94.5] | 47 | 8.1 [5.3-12.1] | 628 | 100 |
| Age | | | | | | | | | | | | |
| 18-29 | 163 | 95.4 [89.9-98.0] | 9 | 4.6 [2.0-10.1] | 172 | 100 | 156 | 95.7 [90.1-98.2] | 9 | 4.3 [1.8-9.9] | 165 | 100 |
| 30-44 | 359 | 88 [83.1-91.7] | 46 | 11.7 [8.1-16.6] | 407 | 100 | 351 | 90.3 [85.7-93.5] | 40 | 9.6 [6.4-14.1] | 392 | 100 |
| 45-59 | 483 | 87.4 [82.6-91.0] | 61 | 12.6 [9.0-17.4] | 544 | 100 | 505 | 88.6 [84.2-91.9] | 59 | 11.4 [8.1-15.8] | 564 | 100 |
| 60+ | 876 | 91.2 [87.5-93.9] | 63 | 7.9 [5.3-11.6] | 946 | 100 | 872 | 89.8 [85.8-92.7] | 68 | 9.1 [6.3-13.0] | 948 | 100 |
| Household gun ownership | | | | | | | | | | | | |

| | | | | | | | | | | | | |
|---|---|---|---|---|---|---|---|---|---|---|---|---|
| No guns in home | 1349 | 90.1 [87.5-92.2] | 123 | 9.7 [7.6-12.3] | 1,478 | 100 | 1324 | 90.4 [87.9-92.4] | 124 | 9.4 [7.4-11.9] | 1,454 | 100 |
| Gun owner | 301 | 89.4 [83.0-93.5] | 29 | 9.7 [5.7-16.0] | 333 | 100 | 326 | 91.4 [85.5-95.0] | 26 | 7.7 [4.3-13.5] | 355 | 100 |
| Non-owner living with owner | 149 | 88.7 [77.8-94.6] | 18 | 11.3 [5.4-22.2] | 167 | 100 | 147 | 89.7 [78.0-95.5] | 16 | 10.3 [4.5-22.0] | 163 | 100 |
| Acquired firearms due to the coronavirus pandemic | | | | | | | | | | | | |
| Non-gun owner | 1580 | 90.1 [87.8-92.1] | 150 | 9.7 [7.7-12.1] | 1,736 | 100 | 1558 | 90.6 [88.3-92.4] | 150 | 9.3 [7.4-11.5] | 1,714 | 100 |
| Gun owner, no purchase | 297 | 89.6 [83.1-93.8] | 27 | 9.4 [5.4-15.9] | 327 | 100 | 320 | 91.6 [85.6-95.2] | 24 | 7.5 [4.1-13.5] | 347 | 100 |
| Gun owner, purchase | 4 | 76 [33.7-95.2] | 2 | 24 [4.8-66.3] | 6 | 100 | 6 | 80.5 [42.4-95.8] | 2 | 19.5 [4.2-57.6] | 8 | 100 |

See footnotes at the end of eAppendix 17.

eAppendix 4. Percentage of respondents willing to ask for help from a hotline, among those worried about intimate partner violence happening to them, California Safety and Wellbeing Survey 2020

| | Not at all willing | | Somewhat willing | | Very willing | | Total | | Somewhat vs. not all willing | Very vs. not at all willing |
|---|---|---|---|---|---|---|---|---|---|---|
| | n | % [95% CI] | n | % [95% CI] | n | % [95% CI] | n | % | PD | PD |
| Total | 50 | 24.6 [16.7-34.5] | 68 | 33.8 [25.3-43.5] | 91 | 41.7 [32.1-51.9] | 209 | 100 | 9.2 | 17.1[a] |
| Education | | | | | | | | | | |
| Less than high school | 5 | 20.5 [8.1-43.1] | 10 | 30.6 [15.8-51.0] | 16 | 48.8 [29.4-68.7] | 31 | 100 | 10.1 | 28.3 |
| High school | 7 | 27.4 [12.3-50.4] | 8 | 33.6 [16.8-56.0] | 14 | 39 [21.5-60.0] | 29 | 100 | 6.3 | 11.7 |
| Some college | 13 | 26.4 [13.4-45.3] | 18 | 35 [21.0-52.1] | 28 | 38.7 [24.8-54.7] | 59 | 100 | 8.6 | 12.3 |
| Bachelor's degree | 17 | 20.8 [9.4-39.9] | 20 | 41.5 [22.5-63.5] | 19 | 37.7 [18.4-61.8] | 56 | 100 | 20.7 | 16.9 |
| Advanced degree | 8 | 37.9 [14.6-68.5] | 12 | 29 [12.7-53.4] | 14 | 33.1 [13.4-61.3] | 34 | 100 | -8.9 | -4.8 |
| Gender | | | | | | | | | | |
| Male | 43 | 30 [20.2-42.1] | 52 | 33.9 [24.2-45.1] | 65 | 36.1 [25.9-47.8] | 160 | 100 | 3.9 | 6.1 |
| Female | 7 | 8.3 [3.3-19.5] | 16 | 33.4 [17.9-53.6] | 26 | 58.3 [38.5-75.7] | 49 | 100 | 25.0[a] | 49.9[a] |
| Race/ethnicity | | | | | | | | | | |
| White, Non-Hispanic | 29 | 37.5 [22.4-55.5] | 18 | 20.3 [10.9-34.7] | 26 | 42.2 [25.7-60.7] | 73 | 100 | -17.2 | 4.7 |
| Black, Non-Hispanic | 2 | 26.7 [6.1-67.0] | 2 | 27.8 [6.3-68.7] | 3 | 45.4 [15.0-79.8] | 7 | 100 | 1.1 | 18.7 |
| Asian, Non-Hispanic | 7 | 43.4 [20.8-69.0] | 8 | 33.5 [14.7-59.5] | 3 | 23.1 [7.2-54.0] | 18 | 100 | -9.8 | -20.3 |
| Other & 2+ races, Non-Hispanic | 0 | 0 [NA] | 1 | 20.8 [2.2-75.0] | 2 | 79.2 [25.0-97.8] | 3 | 100 | 20.8 | 79.2[a] |
| Hispanic | 12 | 11.3 [4.3-26.6] | 39 | 41.4 [28.5-55.5] | 57 | 47.3 [33.6-61.5] | 108 | 100 | 30.1[a] | 36.0[a] |
| Income | | | | | | | | | | |
| Less than $25,000 | 7 | 17.6 [5.1-45.5] | 12 | 27.5 [12.7-49.8] | 25 | 54.9 [31.5-76.3] | 44 | 100 | 10.0 | 37.4[b] |
| $25,000-$59,999 | 14 | 13.5 [6.5-26.1] | 22 | 39.6 [24.2-57.3] | 27 | 46.9 [30.7-63.7] | 63 | 100 | 26.1[a] | 33.3[a] |
| $60,000-$99,999 | 11 | 24.9 [11.9-44.9] | 14 | 36.8 [19.8-58.0] | 20 | 38.3 [21.7-58.2] | 45 | 100 | 12.0 | 13.4 |
| $100,000 or more | 18 | 38.2 [21.7-58.1] | 20 | 30.8 [17.3-48.6] | 19 | 31 [16.0-51.4] | 57 | 100 | -7.4 | -7.3 |
| Age | | | | | | | | | | |
| 18-29 | 1 | 3.9 [0.5-25.3] | 5 | 31.6 [9.4-67.1] | 4 | 64.6 [29.5-88.8] | 10 | 100 | 27.7[b] | 60.7[a] |
| 30-44 | 8 | 21.6 [9.7-41.2] | 22 | 36.4 [21.4-54.7] | 19 | 42 [25.1-61.0] | 49 | 100 | 14.8 | 20.5 |
| 45-59 | 19 | 31.7 [17.5-50.4] | 21 | 36.6 [22.5-53.3] | 30 | 31.7 [19.6-46.9] | 70 | 100 | 4.8 | 0.0 |
| 60+ | 22 | 26 [14.3-42.6] | 20 | 26.4 [14.3-43.5] | 38 | 47.6 [30.6-65.1] | 80 | 100 | 0.3 | 21.5 |

| | | | | | | | | | | |
|---|---|---|---|---|---|---|---|---|---|---|
| Household gun ownership | | | | | | | | | | |
| No guns in home | 33 | 25 [15.9-37.1] | 49 | 35.4 [25.3-47.1] | 65 | 39.6 [28.8-51.4] | 147 | 100 | 10.4 | 14.6 |
| Gun owner | 10 | 19.3 [8.4-38.3] | 10 | 36.9 [16.8-63.0] | 11 | 43.8 [19.8-71.1] | 31 | 100 | 17.6 | 24.5 |
| Non-owner living with owner | 3 | 22.7 [4.1-67.0] | 7 | 22.8 [7.6-51.7] | 10 | 54.5 [21.7-83.7] | 20 | 100 | 0.1 | 31.8 |
| Acquired firearms due to the coronavirus pandemic | | | | | | | | | | |
| Non-gun owner | 40 | 25.3 [16.7-36.4] | 58 | 33.3 [24.3-43.8] | 80 | 41.4 [31.2-52.4] | 178 | 100 | 8.1 | 16.1 |
| Gun owner, no purchase | 8 | 15.7 [6.2-34.3] | 10 | 38.6 [17.3-65.3] | 11 | 45.8 [20.8-73.1] | 29 | 100 | 22.9 | 30.1 |
| Gun owner, purchase | 2 | 100 [NA] | 0 | 0 [NA] | 0 | 0 [NA] | 2 | 100 | -100.0 | -100.0 |

See footnotes at the end of eAppendix 17.

eAppendix 5. Percentage of respondents willing to ask for help from family, among those worried about intimate partner violence happening to them, California Safety and Wellbeing Survey 2020

| | Not at all willing | | Somewhat willing | | Very willing | | Total | | Somewhat vs. not all willing | Very vs. not at all willing |
|---|---|---|---|---|---|---|---|---|---|---|
| | n | % [95% CI] | n | % [95% CI] | n | % [95% CI] | n | % [95% CI] | PD | PD |
| Total | 20 | 13.2 [7.2-22.9] | 75 | 35.3 [26.5-45.1] | 114 | 51.6 [41.5-61.5] | 209 | 100 | 22.1[a] | 38.4[a] |
| Education | | | | | | | | | | |
| Less than high school | 6 | 23.7 [10.1-46.0] | 10 | 33.5 [17.8-53.9] | 15 | 42.8 [24.3-63.6] | 31 | 100 | 9.8 | 19.2 |
| High school | 3 | 10.2 [2.4-35.0] | 11 | 41.1 [22.8-62.3] | 15 | 48.7 [28.8-69.0] | 29 | 100 | 30.9[a] | 38.5[a] |
| Some college | 3 | 5.1 [1.4-16.5] | 21 | 39.5 [24.5-56.9] | 35 | 55.4 [38.6-71.0] | 59 | 100 | 34.4[a] | 50.3[a] |
| Bachelor's degree | 6 | 11.7 [3.4-33.4] | 22 | 27.3 [14.7-44.9] | 28 | 61 [40.5-78.3] | 56 | 100 | 15.6 | 49.3[a] |
| Advanced degree | 2 | 0.9 [0.2-3.8] | 11 | 37.3 [14.4-67.8] | 21 | 61.8 [31.8-84.9] | 34 | 100 | 36.4[a] | 61.0[a] |
| Gender | | | | | | | | | 0.0 | 0.0 |
| Male | 15 | 14.4 [7.1-26.9] | 60 | 36.7 [26.6-48.1] | 85 | 48.9 [37.6-60.4] | 160 | 100 | 22.3[a] | 34.6[a] |
| Female | 5 | 9.7 [3.4-24.8] | 15 | 31 [16.0-51.3] | 29 | 59.4 [39.3-76.7] | 49 | 100 | 21.3[b] | 49.7[a] |
| Race/ethnicity | | | | | | | | | | |
| White, Non-Hispanic | 5 | 11.9 [3.1-36.0] | 27 | 27.9 [16.1-43.9] | 41 | 60.2 [42.2-75.8] | 73 | 100 | 16.0 | 48.3[a] |
| Black, Non-Hispanic | 0 | 0 [NA] | 3 | 33.7 [9.7-70.5] | 4 | 66.3 [29.5-90.3] | 7 | 100 | 33.7[b] | 66.3[a] |
| Asian, Non-Hispanic | 2 | 9.9 [2.3-34.2] | 8 | 38.7 [17.7-65.0] | 8 | 51.4 [26.9-75.2] | 18 | 100 | 28.8[b] | 41.4[a] |
| Other & 2+ races, Non-Hispanic | 0 | 0 [NA] | 1 | 20.8 [2.2-75.0] | 2 | 79.2 [25.0-97.8] | 3 | 100 | 20.8 | 79.2[a] |
| Hispanic | 13 | 15.9 [7.3-31.1] | 36 | 38.1 [25.7-52.4] | 59 | 46 [32.4-60.2] | 108 | 100 | 22.3[a] | 30.1[a] |
| Income | | | | | | | | | | |
| Less than $25,000 | 7 | 30.3 [11.6-58.9] | 13 | 32 [15.2-55.3] | 24 | 37.7 [18.3-62.1] | 44 | 100 | 1.7 | 7.4 |
| $25,000-$59,999 | 4 | 9.8 [2.5-31.0] | 24 | 41.7 [26.1-59.2] | 35 | 48.5 [32.3-65.1] | 63 | 100 | 31.9[a] | 38.8[a] |
| $60,000-$99,999 | 4 | 9 [2.8-25.8] | 18 | 35.3 [19.0-55.9] | 23 | 55.7 [36.0-73.7] | 45 | 100 | 26.3[a] | 46.6[a] |
| $100,000 or more | 5 | 8 [2.5-23.0] | 20 | 31.6 [17.5-50.2] | 32 | 60.4 [41.8-76.4] | 57 | 100 | 23.6[a] | 52.4[a] |
| Age | | | | | | | | | | |
| 18-29 | 1 | 29.6 [4.8-77.7] | 4 | 29.5 [8.4-65.5] | 5 | 40.9 [12.6-76.9] | 10 | 100 | -0.1 | 11.3 |
| 30-44 | 7 | 17.8 [7.2-37.7] | 16 | 30.7 [16.7-49.5] | 26 | 51.5 [33.5-69.2] | 49 | 100 | 12.9 | 33.7[a] |
| 45-59 | 7 | 9 [3.2-22.8] | 27 | 42 [27.0-58.7] | 36 | 49 [33.1-65.1] | 70 | 100 | 33.1[a] | 40.0[a] |
| 60+ | 5 | 6.4 [1.8-20.1] | 28 | 34.4 [20.5-51.6] | 47 | 59.2 [41.9-74.5] | 80 | 100 | 28.0[a] | 52.9[a] |

| | | | | | | | | | | |
|---|---|---|---|---|---|---|---|---|---|---|
| Household gun ownership | | | | | | | | | | |
| No guns in home | 15 | 15.8 [8.3-28.3] | 50 | 34 [24.1-45.6] | 82 | 50.2 [38.6-61.7] | 147 | 100 | 18.1[a] | 34.3[a] |
| Gun owner | 1 | 5.5 [0.8-31.1] | 14 | 40.6 [19.4-66.0] | 16 | 53.9 [28.8-77.2] | 31 | 100 | 35.0[a] | 48.4[a] |
| Non-owner living with owner | 1 | 0.7 [0.1-5.3] | 8 | 39.8 [13.1-74.4] | 11 | 59.5 [25.2-86.5] | 20 | 100 | 39.1[a] | 58.9[a] |
| Acquired firearms due to the coronavirus pandemic | | | | | | | | | | |
| Non-gun owner | 19 | 14.2 [7.6-25.1] | 61 | 34.5 [25.2-45.2] | 98 | 51.3 [40.5-61.9] | 178 | 100 | 20.3[a] | 37.0[a] |
| Gun owner, no purchase | 1 | 5.8 [0.8-32.2] | 13 | 39.6 [18.2-66.0] | 15 | 54.6 [28.6-78.3] | 29 | 100 | 33.8[a] | 48.8[a] |
| Gun owner, purchase | 0 | 0 [NA] | 1 | 61.7 [9.1-96.3] | 1 | 38.3 [3.7-90.9] | 2 | 100 | 61.8[b] | 38.2 |

See footnotes at the end of eAppendix 17.

eAppendix 6. Percentage of respondents willing to ask for help from law enforcement, among those worried about intimate partner violence happening to them, California Safety and Wellbeing Survey 2020

| | Not at all willing | | Somewhat willing | | Very willing | | Total | | Somewhat vs. not all willing | Very vs. not at all willing |
|---|---|---|---|---|---|---|---|---|---|---|
| | n | % [95% CI] | n | % [95% CI] | n | % [95% CI] | n | % [95% CI] | PD | PD |
| Total | 22 | 10.5 [5.4-19.5] | 85 | 42.9 [33.4-52.9] | 102 | 46.6 [36.8-56.7] | 209 | 100 | 32.4[a] | 36.1[a] |
| Education | | | | | | | | | | |
| Less than high school | 4 | 16.3 [5.5-39.7] | 14 | 44.2 [25.8-64.3] | 13 | 39.4 [21.6-60.7] | 31 | 100 | 27.9[b] | 23.1 |
| High school | 1 | 6.1 [0.9-32.6] | 9 | 38 [20.1-59.9] | 19 | 55.9 [34.7-75.1] | 29 | 100 | 31.9[a] | 49.8[a] |
| Some college | 3 | 6.7 [2.1-19.6] | 22 | 31.8 [19.0-48.1] | 34 | 61.4 [45.0-75.6] | 59 | 100 | 25.1[a] | 54.7[a] |
| Bachelor's degree | 10 | 8.4 [3.6-18.3] | 25 | 52.1 [30.6-72.8] | 21 | 39.5 [20.0-63.0] | 56 | 100 | 43.6[a] | 31.1[a] |
| Advanced degree | 4 | 7.7 [1.7-28.8] | 15 | 56 [29.5-79.4] | 15 | 36.3 [15.5-63.9] | 34 | 100 | 48.2[a] | 28.6[b] |
| Gender | | | | | | | | | | |
| Male | 15 | 10.9 [4.8-23.0] | 66 | 43.7 [32.8-55.3] | 79 | 45.4 [34.4-56.9] | 160 | 100 | 32.8[a] | 34.5[a] |
| Female | 7 | 9.3 [3.9-20.8] | 19 | 40.5 [23.2-60.5] | 23 | 50.2 [30.7-69.7] | 49 | 100 | 31.1[a] | 40.9[a] |
| Race/ethnicity | | | | | | | | | | |
| White, Non-Hispanic | 10 | 18.4 [7.3-39.1] | 26 | 26.7 [15.4-42.1] | 37 | 55 [37.4-71.4] | 73 | 100 | 8.3 | 36.6[a] |
| Black, Non-Hispanic | 0 | 0 [NA] | 3 | 53.7 [20.0-84.3] | 4 | 46.3 [15.7-80.0] | 7 | 100 | 53.7[a] | 46.3[a] |
| Asian, Non-Hispanic | 1 | 3.5 [0.5-21.9] | 10 | 51.7 [27.1-75.6] | 7 | 44.7 [21.7-70.3] | 18 | 100 | 48.2[a] | 41.2[a] |
| Other & 2+ races, Non-Hispanic | 0 | 0 [NA] | 2 | 72.6 [18.3-96.9] | 1 | 27.4 [3.1-81.7] | 3 | 100 | 72.6[a] | 27.4 |
| Hispanic | 11 | 9.6 [3.4-24.4] | 44 | 47.2 [33.5-61.3] | 53 | 43.2 [30.0-57.5] | 108 | 100 | 37.6[a] | 33.6[a] |
| Income | | | | | | | | | | |
| Less than $25,000 | 4 | 14 [3.1-45.4] | 15 | 36.2 [18.1-59.3] | 25 | 49.8 [27.0-72.7] | 44 | 100 | 22.2 | 35.8[b] |
| $25,000-$59,999 | 3 | 2.2 [0.6-8.2] | 28 | 52.3 [35.7-68.4] | 32 | 45.5 [29.8-62.1] | 63 | 100 | 50.1[a] | 43.2[a] |
| $60,000-$99,999 | 8 | 16.2 [6.8-33.9] | 17 | 36.2 [19.7-56.7] | 20 | 47.6 [28.9-67.0] | 45 | 100 | 20.0 | 31.3[a] |
| $100,000 or more | 7 | 12.5 [3.7-34.7] | 25 | 42.3 [25.7-60.9] | 25 | 45.2 [27.6-64.1] | 57 | 100 | 29.8[a] | 32.7[a] |
| Age | | | | | | | | | | |
| 18-29 | 2 | 6 [1.2-24.4] | 3 | 25.6 [6.6-62.6] | 5 | 68.4 [32.9-90.6] | 10 | 100 | 19.7 | 62.5[a] |
| 30-44 | 4 | 10.3 [3.0-29.8] | 25 | 50.8 [32.8-68.6] | 20 | 38.9 [22.7-57.9] | 49 | 100 | 40.6[a] | 28.6[a] |
| 45-59 | 7 | 11.3 [3.1-33.6] | 27 | 44.3 [29.0-60.8] | 36 | 44.4 [29.3-60.5] | 70 | 100 | 33.0[a] | 33.0[a] |
| 60+ | 9 | 11.2 [4.3-26.0] | 30 | 34.7 [20.9-51.8] | 41 | 54.1 [36.9-70.4] | 80 | 100 | 23.5[a] | 42.9[a] |

| | | | | | | | | | | |
|---|---|---|---|---|---|---|---|---|---|---|
| Household gun ownership | | | | | | | | | | |
| No guns in home | 16 | 11.3 [5.1-23.1] | 65 | 46.8 [35.5-58.5] | 66 | 41.9 [30.9-53.7] | 147 | 100 | 35.5[a] | 30.6[a] |
| Gun owner | 3 | 8.5 [2.0-30.0] | 8 | 23.6 [8.8-49.8] | 20 | 67.9 [42.1-86.0] | 31 | 100 | 15.2 | 59.5[a] |
| Non-owner living with owner | 2 | 8.5 [1.5-35.5] | 11 | 48.2 [17.7-80.1] | 7 | 43.4 [13.9-78.4] | 20 | 100 | 39.7[b] | 34.9 |
| Acquired firearms due to the coronavirus pandemic | | | | | | | | | | |
| Non-gun owner | 19 | 10.8 [5.2-20.9] | 77 | 45.5 [35.1-56.3] | 82 | 43.7 [33.4-54.7] | 178 | 100 | 34.7 | 33.0 |
| Gun owner, no purchase | 3 | 8.8 [2.0-31.2] | 8 | 24.7 [9.1-51.7] | 18 | 66.5 [39.9-85.6] | 29 | 100 | 15.9 | 57.6 |
| Gun owner, purchase | 0 | 0 [NA] | 0 | 0 [NA] | 2 | 100 [NA] | 2 | 100 | 0.0 | 100.0 |

See footnotes at the end of eAppendix 17.

eAppendix 7. Percentage of respondents worried about homicide happening to them before and during the coronavirus pandemic, California Safety and Wellbeing Survey 2020

| | Before the pandemic | | | | | | During the pandemic | | | | | | Total | | During - Before | | |
|---|---|---|---|---|---|---|---|---|---|---|---|---|---|---|---|---|---|
| | Not worried | | Somewhat worried | | Very worried | | Not worried | | Somewhat worried | | Very worried | | | | Not | Somewhat | Very |
| | n | % [95% CI] | n | % [95% CI] | n | % [95% CI] | n | % [95% CI] | n | % [95% CI] | n | % [95% CI] | n | % | PD | PD | PD |
| Total | 1821 | 54.7 [51.8-57.5] | 784 | 32.1 [29.5-34.8] | 234 | 11.9 [10.1-14.1] | 1728 | 51.2 [48.4-54.0] | 792 | 32.5 [29.9-35.2] | 320 | 15.1 [13.1-17.4] | 2,870 | 100 | -3.5[b] | 0.4 | 3.2[a] |
| Education | | | | | | | | | | | | | | | | | |
| Less than high school | 46 | 28 [20.9-36.5] | 66 | 44.6 [35.9-53.6] | 43 | 24.9 [18.0-33.4] | 47 | 29 [21.7-37.6] | 59 | 39.4 [31.1-48.5] | 46 | 28.5 [21.1-37.2] | 158 | 100 | 0.9 | -5.1 | 3.6 |
| High school | 167 | 46.6 [39.9-53.4] | 113 | 31.5 [25.4-38.2] | 73 | 20 [15.0-26.2] | 153 | 40.7 [34.2-47.5] | 117 | 34.8 [28.5-41.7] | 87 | 23 [17.7-29.3] | 364 | 100 | -5.9 | 3.4 | 2.9 |
| Some college | 601 | 61.5 [57.0-65.8] | 265 | 29.8 [25.9-34.1] | 65 | 7.8 [5.6-10.8] | 576 | 59 [54.5-63.3] | 263 | 29.8 [25.9-34.1] | 93 | 10.7 [8.2-13.9] | 938 | 100 | -2.5 | 0.0 | 2.9 |
| Bachelor's degree | 547 | 63.4 [57.7-68.8] | 190 | 30.2 [25.2-35.7] | 34 | 5.5 [3.0-9.8] | 509 | 58.6 [52.9-64.1] | 203 | 30.9 [25.8-36.4] | 58 | 9.7 [6.6-14.2] | 775 | 100 | -4.8 | 0.7 | 4.3[b] |
| Advanced degree | 460 | 69 [62.5-74.8] | 150 | 27 [21.5-33.4] | 19 | 3.6 [1.8-7.2] | 443 | 63.5 [56.7-69.7] | 150 | 29.6 [23.6-36.5] | 36 | 6 [3.7-9.7] | 635 | 100 | -5.5 | 2.6 | 2.4 |
| Gender | | | | | | | | | | | | | | | | | |
| Male | 988 | 56 [51.9-60.1] | 381 | 30.1 [26.4-34.1] | 121 | 12.7 [9.9-16.2] | 944 | 52.7 [48.6-56.8] | 399 | 32.2 [28.4-36.3] | 148 | 13.8 [11.1-17.2] | 1,506 | 100 | -3.3 | 2.1 | 1.1 |
| Female | 833 | 53.4 [49.6-57.2] | 403 | 33.9 [30.3-37.7] | 113 | 11.2 [8.8-14.1] | 784 | 49.7 [45.9-53.5] | 393 | 32.7 [29.2-36.4] | 172 | 16.3 [13.5-19.6] | 1,364 | 100 | -3.7 | -1.2 | 5.1[a] |
| Race/ethnicity | | | | | | | | | | | | | | | | | |
| White, Non-Hispanic | 1226 | 73.7 [70.2-76.9] | 322 | 20.2 [17.4-23.2] | 55 | 5.3 [3.6-7.8] | 1169 | 68.2 [64.5-71.7] | 364 | 25.2 [22.1-28.7] | 72 | 5.8 [4.1-8.1] | 1,615 | 100 | -5.5[a] | 5.1[a] | 0.5 |
| Black, Non-Hispanic | 65 | 43.2 [32.0-55.1] | 48 | 44.9 [33.2-57.1] | 13 | 11.8 [6.0-22.0] | 66 | 44.8 [33.4-56.8] | 48 | 43.4 [31.8-55.7] | 12 | 11.2 [5.6-21.4] | 127 | 100 | 1.7 | -1.5 | -0.6 |
| Asian, Non-Hispanic | 103 | 48.9 [40.5-57.4] | 86 | 41.8 [33.7-50.4] | 19 | 9.3 [5.3-15.7] | 102 | 44.7 [36.5-53.2] | 73 | 39.9 [31.8-48.6] | 33 | 15.4 [10.2-22.6] | 208 | 100 | -4.2 | -2.0 | 6.1 |
| Other & 2+ races, Non-Hispanic | 47 | 62.4 [44.7-77.3] | 20 | 31.7 [17.8-50.0] | 3 | 3.6 [1.0-12.5] | 50 | 68.3 [50.3-82.1] | 16 | 22.4 [11.4-39.1] | 5 | 9.3 [2.5-29.5] | 71 | 100 | 5.9 | -9.4 | 5.7 |
| Hispanic | 380 | 35.3 [30.9-40.0] | 308 | 40.4 [35.5-45.4] | 144 | 21.9 [17.8-26.6] | 341 | 32.7 [28.4-37.4] | 291 | 37.3 [32.6-42.2] | 198 | 27.5 [23.1-32.3] | 849 | 100 | -2.6 | -3.1 | 5.6[b] |
| Income | | | | | | | | | | | | | | | | | |
| Less than $25,000 | 188 | 34.4 [28.2-41.2] | 149 | 40.4 [33.5-47.7] | 84 | 21.6 [16.2-28.1] | 199 | 39.7 [33.0-46.8] | 124 | 34.3 [27.8-41.5] | 96 | 22.4 [16.9-29.1] | 431 | 100 | 5.4 | -6.1 | 0.9 |
| $25,000-$59,999 | 435 | 43.6 [38.4-48.9] | 222 | 38.2 [32.9-43.8] | 79 | 17.6 [13.4-22.7] | 400 | 38.4 [33.5-43.4] | 237 | 39.3 [34.0-44.9] | 101 | 21.6 [17.1-26.9] | 744 | 100 | -5.2 | 1.0 | 4.0 |
| $60,000-$99,999 | 493 | 55.1 [49.4-60.7] | 212 | 33.6 [28.4-39.2] | 46 | 10.2 [6.8-14.9] | 475 | 51.9 [46.3-57.5] | 198 | 31 [25.9-36.6] | 78 | 16.4 [12.3-21.5] | 757 | 100 | -3.2 | -2.6 | 6.2[a] |
| $100,000 or more | 705 | 69.2 [64.5-73.6] | 201 | 24.1 [20.2-28.5] | 25 | 5.7 [3.5-9.2] | 654 | 63.2 [58.3-67.7] | 233 | 28.4 [24.2-33.0] | 45 | 7.5 [5.0-11.0] | 938 | 100 | -6.1[b] | 4.3 | 1.8 |
| Age | | | | | | | | | | | | | | | | | |
| 18-29 | 105 | 51.3 [42.9-59.6] | 71 | 35.1 [27.5-43.5] | 21 | 12.4 [7.6-19.4] | 102 | 47.9 [39.6-56.3] | 63 | 35.3 [27.6-43.8] | 33 | 16 [10.7-23.3] | 200 | 100 | -3.4 | 0.2 | 3.6 |

| | | | | | | | | | | | | | | | | | |
|---|---|---|---|---|---|---|---|---|---|---|---|---|---|---|---|---|---|
| 30-44 | 248 | 47.3 [41.5-53.1] | 187 | 35.2 [29.9-40.9] | 69 | 16.3 [12.2-21.5] | 231 | 42.7 [37.0-48.5] | 181 | 37.4 [31.9-43.2] | 91 | 18.6 [14.4-23.7] | 511 | 100 | -4.6 | 2.2 | 2.3 |
| 45-59 | 428 | 51 [45.9-56.1] | 225 | 34.8 [30.0-40.0] | 76 | 12.1 [9.1-16.1] | 394 | 48.1 [43.1-53.2] | 231 | 33.2 [28.5-38.2] | 104 | 17 [13.3-21.5] | 738 | 100 | -2.9 | -1.6 | 4.9[b] |
| 60+ | 1040 | 68.3 [64.4-71.9] | 301 | 24.2 [20.9-27.9] | 68 | 6.8 [4.8-9.4] | 1001 | 65.4 [61.5-69.0] | 317 | 24.8 [21.6-28.3] | 92 | 9 [6.7-12.0] | 1,421 | 100 | -2.9 | 0.6 | 2.2 |
| Household gun ownership | | | | | | | | | | | | | | | | | |
| No guns in home | 1208 | 51.2 [47.9-54.6] | 570 | 34.2 [31.0-37.5] | 192 | 13.4 [11.1-16.0] | 1154 | 48 [44.7-51.3] | 563 | 34 [30.8-37.3] | 255 | 16.8 [14.3-19.7] | 1,989 | 100 | -3.2 | -0.2 | 3.5[b] |
| Gun owner | 387 | 68.4 [61.6-74.4] | 124 | 25.8 [20.3-32.1] | 16 | 5.3 [2.6-10.6] | 361 | 64 [57.1-70.3] | 138 | 28 [22.2-34.7] | 28 | 7.9 [4.7-13.2] | 529 | 100 | -4.4 | 2.2 | 2.7 |
| Non-owner living with owner | 149 | 61.2 [51.1-70.5] | 57 | 26.5 [18.9-35.9] | 12 | 12 [6.1-22.4] | 142 | 59.1 [49.1-68.4] | 52 | 24.4 [17.1-33.5] | 24 | 16.2 [9.5-26.2] | 219 | 100 | -2.1 | -2.1 | 4.1 |
| Acquired firearms due to the coronavirus pandemic | | | | | | | | | | | | | | | | | |
| Non-gun owner | 1434 | 52.2 [49.1-55.3] | 660 | 33.2 [30.3-36.2] | 218 | 13.1 [11.0-15.6] | 1367 | 48.9 [45.8-51.9] | 654 | 33.3 [30.4-36.3] | 292 | 16.4 [14.1-19.0] | 2,341 | 100 | -3.3 | 0.1 | 3.3 |
| Gun owner, no purchase | 378 | 67.7 [60.8-73.9] | 123 | 26.3 [20.7-32.8] | 16 | 5.4 [2.6-10.9] | 354 | 63.5 [56.4-70.0] | 137 | 28.6 [22.7-35.4] | 26 | 7.8 [4.5-13.2] | 519 | 100 | -4.2 | 2.3 | 2.4 |
| Gun owner, purchase | 9 | 95.5 [72.0-99.4] | 1 | 4.5 [0.6-28.0] | 0 | 0 [NA] | 7 | 84.6 [56.0-96.0] | 1 | 3.6 [0.5-23.7] | 2 | 11.7 [2.5-40.9] | 10 | 100 | -10.9 | -0.9 | 11.7 |

See footnotes at the end of eAppendix 17.

eAppendix 8. Percentage of respondents worried about suicide happening to them before and during the coronavirus pandemic, California Safety and Wellbeing Survey 2020

| | Before the pandemic | | | | | | During the pandemic | | | | | | Total | | During - Before | | |
|---|---|---|---|---|---|---|---|---|---|---|---|---|---|---|---|---|---|
| | Not worried | | Somewhat worried | | Very worried | | Not worried | | Somewhat worried | | Very worried | | | | Not | Somewhat | Very |
| | n | % [95% CI] | n | % [95% CI] | n | % [95% CI] | n | % [95% CI] | n | % [95% CI] | n | % [95% CI] | n | % | PD | PD | PD |
| Total | 2351 | 74.6 [71.9-77.1] | 335 | 16.6 [14.5-19.1] | 162 | 7.9 [6.4-9.7] | 2293 | 71 [68.3-73.7] | 353 | 18.7 [16.4-21.2] | 199 | 9.1 [7.5-10.9] | 2,870 | 100 | -3.5[b] | 2.1 | 1.2 |
| Education | | | | | | | | | | | | | | | | | |
| Less than high school | 79 | 52.2 [43.3-61.0] | 45 | 31 [23.3-39.9] | 31 | 15.1 [9.9-22.3] | 70 | 43.9 [35.4-52.9] | 51 | 36.3 [28.1-45.3] | 31 | 16.7 [11.1-24.4] | 158 | 100 | -8.3 | 5.3 | 1.6 |
| High school | 257 | 70.2 [63.6-76.1] | 55 | 17 [12.4-22.9] | 45 | 11.4 [7.8-16.5] | 241 | 65.9 [59.1-72.0] | 62 | 19.9 [14.9-26.1] | 53 | 12.5 [8.6-17.6] | 364 | 100 | -4.4 | 2.9 | 1.0 |
| Some college | 781 | 78.4 [73.9-82.3] | 110 | 14.5 [11.2-18.5] | 42 | 6.5 [4.4-9.6] | 775 | 78 [73.7-81.8] | 104 | 14.6 [11.3-18.6] | 54 | 6.8 [4.8-9.5] | 938 | 100 | -0.4 | 0.1 | 0.2 |
| Bachelor's degree | 664 | 80.6 [75.3-85.0] | 80 | 14 [10.2-19.0] | 30 | 4.9 [2.9-8.1] | 650 | 77.1 [71.6-81.8] | 82 | 14.1 [10.5-18.6] | 41 | 8.3 [5.2-13.0] | 775 | 100 | -3.5 | 0.1 | 3.4 |
| Advanced degree | 570 | 89 [84.1-92.5] | 45 | 8.5 [5.3-13.4] | 14 | 1.8 [0.9-3.5] | 557 | 84.6 [78.8-89.0] | 54 | 12.8 [8.6-18.7] | 20 | 1.9 [1.1-3.5] | 635 | 100 | -4.4 | 4.3 | 0.2 |
| Gender | | | | | | | | | | | | | | | | | |
| Male | 1261 | 75.1 [71.0-78.8] | 163 | 17.2 [13.9-21.0] | 69 | 6.6 [4.8-9.1] | 1224 | 71.9 [67.8-75.7] | 168 | 18 [14.8-21.8] | 101 | 8.6 [6.5-11.3] | 1,506 | 100 | -3.2 | 0.8 | 2.0 |
| Female | 1090 | 74.1 [70.4-77.4] | 172 | 16.1 [13.4-19.3] | 93 | 9 [6.9-11.7] | 1069 | 70.3 [66.4-73.8] | 185 | 19.3 [16.2-22.8] | 98 | 9.5 [7.3-12.2] | 1,364 | 100 | -3.8 | 3.2 | 0.4 |
| Race/ethnicity | | | | | | | | | | | | | | | | | |
| White, Non-Hispanic | 1444 | 84.6 [81.2-87.5] | 125 | 10.8 [8.4-13.8] | 37 | 3.8 [2.4-6.0] | 1434 | 84.8 [81.6-87.5] | 128 | 10.5 [8.3-13.2] | 46 | 4 [2.6-6.2] | 1,615 | 100 | 0.2 | -0.3 | 0.3 |
| Black, Non-Hispanic | 102 | 69.6 [56.3-80.2] | 19 | 22.3 [13.2-35.2] | 6 | 8.2 [3.0-20.4] | 104 | 70.1 [56.7-80.7] | 19 | 24.8 [15.0-38.1] | 4 | 5.1 [1.4-16.3] | 127 | 100 | 0.5 | 2.5 | -3.1 |
| Asian, Non-Hispanic | 162 | 77.3 [69.4-83.7] | 33 | 17 [11.4-24.7] | 12 | 5.3 [2.7-10.2] | 152 | 72.2 [63.8-79.2] | 38 | 19.7 [13.6-27.6] | 18 | 8.2 [4.6-14.1] | 208 | 100 | -5.2 | 2.6 | 2.8 |
| Other & 2+ races, Non-Hispanic | 60 | 84 [67.5-93.0] | 10 | 14.9 [6.2-31.7] | 1 | 1.1 [0.1-7.3] | 57 | 79.7 [63.4-89.9] | 13 | 19.2 [9.3-35.6] | 1 | 1.1 [0.1-7.3] | 71 | 100 | -4.3 | 4.3 | 0.0 |
| Hispanic | 583 | 61.2 [56.1-66.1] | 148 | 22.7 [18.6-27.5] | 106 | 14.5 [11.3-18.5] | 546 | 53.3 [48.2-58.3] | 155 | 27.1 [22.6-32.1] | 130 | 16.9 [13.3-21.1] | 849 | 100 | -7.9[a] | 4.4 | 2.3 |
| Income | | | | | | | | | | | | | | | | | |
| Less than $25,000 | 274 | 54 [46.7-61.1] | 85 | 27.2 [21.0-34.5] | 62 | 15.3 [10.8-21.3] | 267 | 51.8 [44.6-58.9] | 80 | 26.9 [20.8-33.9] | 74 | 17.6 [12.6-23.9] | 431 | 100 | -2.1 | -0.3 | 2.3 |
| $25,000-$59,999 | 597 | 70.6 [65.0-75.7] | 95 | 18.8 [14.6-24.0] | 49 | 10.3 [7.2-14.6] | 575 | 65.3 [59.5-70.6] | 101 | 21.6 [17.0-27.1] | 61 | 11.9 [8.6-16.3] | 744 | 100 | -5.4 | 2.8 | 1.6 |
| $60,000-$99,999 | 636 | 76.3 [70.7-81.1] | 88 | 16.2 [12.1-21.3] | 31 | 7.4 [4.7-11.4] | 628 | 73.7 [68.0-78.8] | 86 | 16.9 [12.7-22.2] | 41 | 9.1 [6.1-13.5] | 757 | 100 | -2.5 | 0.7 | 1.8 |
| $100,000 or more | 844 | 84 [79.7-87.6] | 67 | 11.4 [8.3-15.4] | 20 | 3.8 [2.2-6.5] | 823 | 80.5 [76.0-84.4] | 86 | 14.7 [11.3-18.9] | 23 | 3.9 [2.2-6.7] | 938 | 100 | -3.5 | 3.4 | 0.1 |
| Age | | | | | | | | | | | | | | | | | |
| 18-29 | 137 | 69 [60.8-76.3] | 39 | 20.3 [14.2-28.2] | 21 | 9.8 [6.0-15.5] | 131 | 67.5 [59.3-74.8] | 40 | 17.4 [12.1-24.4] | 26 | 13.8 [8.9-20.8] | 200 | 100 | -1.5 | -2.8 | 4.0 |

| | | | | | | | | | | | | | | | | |
|---|---|---|---|---|---|---|---|---|---|---|---|---|---|---|---|---|
| 30-44 | 356 | 65.3 [59.4-70.7] | 100 | 24 [19.2-29.6] | 48 | 9.5 [6.5-13.7] | 328 | 59.8 [53.9-65.4] | 117 | 28.1 [23.0-33.7] | 57 | 10.7 [7.5-15.0] | 511 | 100 | -5.5 | 4.1 | 1.2 |
| 45-59 | 578 | 73.6 [68.5-78.0] | 97 | 15.9 [12.3-20.4] | 57 | 9.3 [6.7-12.8] | 561 | 67.9 [62.6-72.8] | 103 | 20.9 [16.5-26.1] | 65 | 9.3 [6.8-12.5] | 738 | 100 | -5.7 | 5.0 | 0.0 |
| 60+ | 1280 | 88.9 [86.0-91.2] | 99 | 7.1 [5.5-9.3] | 36 | 3.6 [2.1-6.1] | 1273 | 88.4 [85.5-90.8] | 93 | 7.1 [5.4-9.3] | 51 | 4.2 [2.7-6.6] | 1,421 | 100 | -0.5 | 0.0 | 0.6 |
| Household gun ownership | | | | | | | | | | | | | | | | | |
| No guns in home | 1588 | 71.8 [68.5-74.9] | 252 | 18 [15.4-21.0] | 136 | 9.4 [7.5-11.7] | 1531 | 67.2 [63.8-70.4] | 275 | 21.2 [18.4-24.3] | 166 | 10.4 [8.4-12.8] | 1,989 | 100 | -4.6[a] | 3.2 | 1.0 |
| Gun owner | 487 | 90.5 [85.2-94.0] | 34 | 8.1 [4.8-13.4] | 6 | 1 [0.4-2.6] | 483 | 88.3 [82.5-92.4] | 33 | 8.6 [5.1-14.0] | 12 | 2.9 [1.2-6.8] | 529 | 100 | -2.2 | 0.5 | 1.9 |
| Non-owner living with owner | 180 | 78.6 [68.7-86.0] | 27 | 12.2 [6.9-20.6] | 12 | 9.2 [4.5-18.2] | 183 | 78.9 [69.0-86.2] | 23 | 11.5 [6.4-19.7] | 13 | 9.6 [4.7-18.7] | 219 | 100 | 0.3 | -0.7 | 0.4 |
| Acquired firearms due to the coronavirus pandemic | | | | | | | | | | | | | | | | | |
| Non-gun owner | 1864 | 71.7 [68.7-74.5] | 301 | 18.2 [15.7-20.9] | 156 | 9.1 [7.4-11.2] | 1810 | 67.9 [64.9-70.9] | 320 | 20.5 [17.9-23.3] | 187 | 10.2 [8.3-12.3] | 2,341 | 100 | -3.8 | 2.3 | 1.0 |
| Gun owner, no purchase | 478 | 90.3 [84.9-93.9] | 33 | 8.2 [4.8-13.6] | 6 | 1 [0.4-2.7] | 474 | 88.1 [82.2-92.3] | 33 | 8.8 [5.2-14.3] | 11 | 2.9 [1.2-6.9] | 519 | 100 | -2.2 | 0.6 | 1.8 |
| Gun owner, purchase | 9 | 95.5 [72.0-99.4] | 1 | 4.5 [0.6-28.0] | 0 | 0 [NA] | 9 | 95.5 [72.0-99.4] | 0 | 0 [NA] | 1 | 4.5 [0.6-28.0] | 10 | 100 | 0.0 | -4.5 | 4.5 |

See footnotes at the end of eAppendix 17.

eAppendix 9. Percentage of respondents worried about a mass shooting happening to them before and during the coronavirus pandemic, California Safety and Wellbeing Survey 2020

| | Before the pandemic | | | | | | During the pandemic | | | | | | Total | | During - Before | | |
|---|---|---|---|---|---|---|---|---|---|---|---|---|---|---|---|---|---|
| | Not worried | | Somewhat worried | | Very worried | | Not worried | | Somewhat worried | | Very worried | | | | Not | Somewhat | Very |
| | n | % [95% CI] | n | % [95% CI] | n | % [95% CI] | n | % [95% CI] | n | % [95% CI] | n | % [95% CI] | n | % | PD | PD | PD |
| Total | 1347 | 39.1 [36.5-41.7] | 1086 | 41.2 [38.4-44.0] | 412 | 18.8 [16.6-21.2] | 1471 | 43.2 [40.5-46.0] | 956 | 37.4 [34.7-40.1] | 410 | 18 [15.8-20.3] | 2,870 | 100 | 4.2[a] | -3.8[b] | -0.8 |
| Education | | | | | | | | | | | | | | | | | |
| Less than high school | 40 | 26.7 [19.5-35.2] | 58 | 37.4 [29.3-46.4] | 58 | 34.3 [26.4-43.2] | 37 | 23.4 [16.7-31.7] | 64 | 41.1 [32.7-50.1] | 54 | 32.2 [24.5-41.0] | 158 | 100 | -3.3 | 3.7 | -2.1 |
| High school | 131 | 34.9 [28.8-41.6] | 133 | 40 [33.4-47.0] | 93 | 23.5 [18.2-29.8] | 128 | 36.5 [30.2-43.4] | 126 | 35.5 [29.3-42.4] | 103 | 26.3 [20.8-32.7] | 364 | 100 | 1.6 | -4.5 | 2.8 |
| Some college | 472 | 46 [41.6-50.4] | 340 | 39.8 [35.4-44.3] | 119 | 13.6 [10.8-16.9] | 479 | 47.5 [43.0-51.9] | 328 | 37.6 [33.4-42.1] | 121 | 13.9 [11.0-17.4] | 938 | 100 | 1.5 | -2.1 | 0.3 |
| Bachelor's degree | 373 | 37.9 [32.9-43.1] | 317 | 47.3 [41.7-52.9] | 80 | 14 [10.2-18.9] | 426 | 47.3 [41.8-52.8] | 262 | 39.8 [34.3-45.4] | 80 | 12.1 [8.7-16.7] | 775 | 100 | 9.4[a] | -7.5[b] | -1.8 |
| Advanced degree | 331 | 44.1 [38.0-50.4] | 238 | 42.8 [36.4-49.5] | 62 | 12.8 [8.8-18.2] | 401 | 60 [53.3-66.2] | 176 | 32.3 [26.2-39.1] | 52 | 6.9 [4.8-9.9] | 635 | 100 | 15.9[a] | -10.5[a] | -5.9[a] |
| Gender | | | | | | | | | | | | | | | | | |
| Male | 800 | 44.3 [40.4-48.4] | 513 | 37.2 [33.2-41.3] | 181 | 17.4 [14.3-21.1] | 849 | 47.7 [43.7-51.8] | 449 | 33.2 [29.4-37.3] | 192 | 17.2 [14.2-20.8] | 1,506 | 100 | 3.4 | -4.0 | -0.2 |
| Female | 547 | 34.2 [30.8-37.8] | 573 | 44.9 [41.1-48.7] | 231 | 20 [17.0-23.3] | 622 | 39.1 [35.5-42.8] | 507 | 41.2 [37.5-45.0] | 218 | 18.6 [15.7-21.9] | 1,364 | 100 | 4.9[b] | -3.7 | -1.3 |
| Race/ethnicity | | | | | | | | | | | | | | | | | |
| White, Non-Hispanic | 929 | 53.7 [50.0-57.5] | 570 | 37.4 [33.8-41.1] | 108 | 8.2 [6.2-10.7] | 1004 | 58 [54.2-61.7] | 485 | 31.9 [28.5-35.5] | 115 | 9.3 [7.1-12.1] | 1,615 | 100 | 4.3 | -5.5[a] | 1.1 |
| Black, Non-Hispanic | 61 | 45.9 [34.3-58.0] | 42 | 35.9 [25.4-48.1] | 24 | 18.2 [10.4-29.7] | 63 | 42.5 [31.4-54.5] | 46 | 44 [32.4-56.3] | 18 | 13.5 [7.4-23.3] | 127 | 100 | -3.4 | 8.1 | -4.7 |
| Asian, Non-Hispanic | 70 | 26.9 [20.4-34.5] | 96 | 53.3 [44.8-61.5] | 40 | 19.3 [13.5-26.8] | 91 | 40.1 [32.2-48.6] | 81 | 43.9 [35.6-52.5] | 34 | 15.3 [10.1-22.3] | 208 | 100 | 13.2[a] | -9.4 | -4.0 |
| Other & 2+ races, Non-Hispanic | 39 | 48.6 [32.1-65.5] | 22 | 37.8 [22.4-56.1] | 10 | 13.6 [6.3-26.9] | 43 | 56.3 [39.1-72.1] | 19 | 29.5 [16.8-46.3] | 9 | 14.2 [5.4-32.4] | 71 | 100 | 7.7 | -8.3 | 0.7 |
| Hispanic | 248 | 24.4 [20.5-28.7] | 356 | 41.9 [37.1-46.9] | 230 | 31.9 [27.3-36.8] | 270 | 25.6 [21.6-29.9] | 325 | 40.9 [36.0-45.9] | 234 | 30.7 [26.3-35.5] | 849 | 100 | 1.2 | -1.1 | -1.2 |
| Income | | | | | | | | | | | | | | | | | |
| Less than $25,000 | 158 | 32.2 [26.0-39.1] | 149 | 36.7 [30.0-43.9] | 116 | 27.8 [21.9-34.6] | 163 | 32.6 [26.4-39.5] | 145 | 36 [29.4-43.2] | 115 | 28 [22.0-34.9] | 431 | 100 | 0.4 | -0.6 | 0.2 |
| $25,000-$59,999 | 339 | 34.1 [29.3-39.1] | 268 | 40.4 [35.2-45.8] | 131 | 25 [20.2-30.5] | 349 | 34.3 [29.6-39.3] | 253 | 40.8 [35.5-46.4] | 129 | 23.3 [18.9-28.5] | 744 | 100 | 0.2 | 0.4 | -1.7 |
| $60,000-$99,999 | 363 | 39.3 [34.1-44.8] | 300 | 41 [35.6-46.7] | 91 | 19.3 [14.9-24.6] | 415 | 43.8 [38.4-49.3] | 237 | 35.4 [30.2-41.0] | 98 | 19.6 [15.2-24.9] | 757 | 100 | 4.5 | -5.6 | 0.4 |
| $100,000 or more | 487 | 44.7 [40.1-49.3] | 369 | 43.6 [38.9-48.3] | 74 | 11 [8.2-14.7] | 544 | 52.6 [47.8-57.3] | 321 | 36.9 [32.4-41.6] | 68 | 9.7 [7.0-13.3] | 938 | 100 | 7.9[a] | -6.6[a] | -1.3 |
| Age | | | | | | | | | | | | | | | | | |
| 18-29 | 55 | 27.4 [20.6-35.5] | 92 | 47.5 [39.1-55.9] | 51 | 24.3 [17.9-32.1] | 79 | 37.9 [30.2-46.3] | 78 | 41.1 [33.0-49.6] | 40 | 19.9 [14.0-27.6] | 200 | 100 | 10.5[b] | -6.4 | -4.4 |

| | | | | | | | | | | | | | | | | | |
|---|---|---|---|---|---|---|---|---|---|---|---|---|---|---|---|---|---|
| 30-44 | 159 | 32.8 [27.6-38.5] | 241 | 44.6 [38.9-50.4] | 105 | 21.6 [17.1-26.9] | 195 | 37.9 [32.4-43.7] | 194 | 39.4 [33.9-45.3] | 113 | 21.3 [17.0-26.5] | 511 | 100 | 5.1 | -5.1 | -0.3 |
| 45-59 | 309 | 36.9 [32.3-41.8] | 295 | 42 [37.0-47.1] | 128 | 19.8 [16.0-24.3] | 337 | 39.6 [34.9-44.6] | 260 | 37.7 [32.8-42.8] | 132 | 20.2 [16.3-24.7] | 738 | 100 | 2.7 | -4.3 | 0.3 |
| 60+ | 824 | 54.9 [51.1-58.6] | 458 | 33 [29.5-36.7] | 128 | 11.3 [8.9-14.3] | 860 | 55.7 [51.9-59.4] | 424 | 32.6 [29.2-36.3] | 125 | 11 [8.5-14.1] | 1,421 | 100 | 0.8 | -0.4 | -0.3 |
| Household gun ownership | | | | | | | | | | | | | | | | | |
| No guns in home | 845 | 35 [32.0-38.1] | 790 | 42.8 [39.5-46.1] | 341 | 21.5 [18.8-24.5] | 950 | 39.6 [36.5-42.9] | 685 | 38.6 [35.4-41.9] | 331 | 20.2 [17.6-23.1] | 1,989 | 100 | 4.7[a] | -4.2[b] | -1.3 |
| Gun owner | 338 | 57.5 [50.5-64.1] | 163 | 35 [28.7-41.9] | 24 | 7.1 [4.0-12.2] | 346 | 56.6 [49.7-63.3] | 148 | 33.2 [27.0-40.0] | 34 | 10.1 [6.3-15.8] | 529 | 100 | -0.8 | -1.9 | 3.1 |
| Non-owner living with owner | 97 | 36.9 [28.4-46.3] | 93 | 42 [32.7-52.0] | 29 | 21 [13.5-31.2] | 109 | 50.1 [40.4-59.8] | 79 | 33.9 [25.7-43.3] | 28 | 15.8 [9.3-25.4] | 219 | 100 | 13.2[b] | -8.1 | -5.3 |
| Acquired firearms due to the coronavirus pandemic | | | | | | | | | | | | | | | | | |
| Non-gun owner | 1009 | 35.8 [33.0-38.6] | 923 | 42.3 [39.3-45.4] | 388 | 20.8 [18.4-23.6] | 1125 | 40.8 [37.9-43.8] | 808 | 38.1 [35.2-41.2] | 376 | 19.4 [17.0-22.0] | 2,341 | 100 | 5.1[a] | -4.2[b] | -1.5 |
| Gun owner, no purchase | 332 | 57.3 [50.3-64.1] | 159 | 35 [28.6-42.0] | 24 | 7.3 [4.1-12.5] | 341 | 56.9 [49.8-63.6] | 143 | 32.7 [26.4-39.7] | 34 | 10.4 [6.5-16.2] | 519 | 100 | -0.5 | -2.3 | 3.1 |
| Gun owner, purchase | 6 | 63.3 [27.4-88.8] | 4 | 36.7 [11.2-72.6] | 0 | 0 [NA] | 5 | 47.5 [16.6-80.4] | 5 | 52.5 [19.6-83.4] | 0 | 0 [NA] | 10 | 100 | -15.8 | 15.8 | 0.0 |

See footnotes at the end of eAppendix 17.

eAppendix 10. Percentage of respondents worried about assault happening to them before and during the coronavirus pandemic, California Safety and Wellbeing Survey 2020

| | Before the pandemic | | | | | | During the pandemic | | | | | | Total | | During - Before | | |
|---|---|---|---|---|---|---|---|---|---|---|---|---|---|---|---|---|---|
| | Not worried | | Somewhat worried | | Very worried | | Not worried | | Somewhat worried | | Very worried | | | | Not | Somewhat | Very |
| | n | % [95% CI] | n | % [95% CI] | n | % [95% CI] | n | % [95% CI] | n | % [95% CI] | n | % [95% CI] | n | % | PD | PD | PD |
| Total | 1291 | 39.9 [37.3-42.6] | 1264 | 45.8 [43.1-48.6] | 293 | 13.3 [11.4-15.4] | 1193 | 36.5 [33.9-39.2] | 1222 | 43.9 [41.1-46.7] | 433 | 18.6 [16.4-20.9] | 2,870 | 100 | -3.4[b] | -2.0 | 5.3[a] |
| Education | | | | | | | | | | | | | | | | | |
| Less than high school | 38 | 23.7 [17.0-32.1] | 70 | 47.1 [38.4-56.0] | 47 | 26.7 [19.6-35.2] | 40 | 24.2 [17.5-32.4] | 64 | 43.4 [34.8-52.4] | 53 | 31.1 [23.5-39.9] | 158 | 100 | 0.4 | -3.7 | 4.4 |
| High school | 128 | 37.2 [30.8-44.1] | 166 | 45.9 [39.2-52.8] | 67 | 15.9 [11.7-21.4] | 101 | 28.7 [22.8-35.4] | 146 | 41.5 [35.0-48.4] | 110 | 27.6 [22.0-34.0] | 364 | 100 | -8.6[b] | -4.4 | 11.6[a] |
| Some college | 422 | 44 [39.6-48.5] | 419 | 44.7 [40.3-49.2] | 89 | 10.5 [8.0-13.7] | 379 | 40.3 [35.9-44.8] | 423 | 44.8 [40.4-49.3] | 131 | 14.4 [11.5-17.8] | 938 | 100 | -3.8 | 0.1 | 3.8[b] |
| Bachelor's degree | 397 | 45.7 [40.3-51.2] | 318 | 43.1 [37.7-48.7] | 57 | 10.6 [7.1-15.4] | 362 | 39.8 [34.7-45.2] | 324 | 46.4 [40.9-52.0] | 85 | 13 [9.4-17.8] | 775 | 100 | -5.9 | 3.3 | 2.5 |
| Advanced degree | 306 | 44.9 [38.6-51.3] | 291 | 50.3 [43.8-56.7] | 33 | 4.3 [2.8-6.6] | 311 | 48.7 [42.3-55.2] | 265 | 42.4 [36.1-49.0] | 54 | 8.1 [5.8-11.3] | 635 | 100 | 3.8 | -7.9[b] | 3.8[a] |
| Gender | | | | | | | | | | | | | | | | | |
| Male | 753 | 44.7 [40.7-48.7] | 615 | 42.6 [38.6-46.7] | 127 | 11.6 [9.1-14.7] | 664 | 39.2 [35.3-43.2] | 625 | 42 [38.0-46.1] | 204 | 17.5 [14.4-20.9] | 1,506 | 100 | -5.5[b] | -0.5 | 5.9[a] |
| Female | 538 | 35.6 [32.1-39.2] | 649 | 48.8 [45.0-52.6] | 166 | 14.8 [12.1-17.9] | 529 | 34.1 [30.7-37.7] | 597 | 45.6 [41.8-49.4] | 229 | 19.6 [16.6-22.9] | 1,364 | 100 | -1.5 | -3.2 | 4.8[a] |
| Race/ethnicity | | | | | | | | | | | | | | | | | |
| White, Non-Hispanic | 879 | 54.1 [50.3-57.8] | 650 | 39.8 [36.2-43.5] | 74 | 5.2 [3.6-7.3] | 802 | 47.5 [43.7-51.2] | 681 | 43.1 [39.4-46.8] | 120 | 8.5 [6.5-10.9] | 1,615 | 100 | -6.6[a] | 3.3 | 3.3[a] |
| Black, Non-Hispanic | 52 | 30.7 [21.3-42.0] | 63 | 60.8 [49.0-71.5] | 12 | 8.5 [3.8-18.0] | 57 | 36.9 [26.4-48.7] | 58 | 52.9 [41.0-64.5] | 12 | 10.2 [4.8-20.4] | 127 | 100 | 6.2 | -7.9 | 1.7 |
| Asian, Non-Hispanic | 70 | 34 [26.5-42.4] | 111 | 52.9 [44.4-61.3] | 27 | 13.1 [8.3-19.9] | 73 | 34.5 [26.9-43.0] | 99 | 49.7 [41.3-58.2] | 36 | 15.8 [10.5-22.9] | 208 | 100 | 0.5 | -3.2 | 2.7 |
| Other & 2+ races, Non-Hispanic | 40 | 46.6 [30.5-63.4] | 24 | 40.8 [24.7-59.2] | 6 | 9.9 [3.8-23.3] | 40 | 49.1 [32.5-65.8] | 24 | 39.5 [23.9-57.6] | 7 | 11.4 [3.7-30.0] | 71 | 100 | 2.5 | -1.3 | 1.5 |
| Hispanic | 250 | 26.2 [22.1-30.8] | 416 | 48.1 [43.1-53.1] | 174 | 24.2 [20.1-28.9] | 221 | 22.9 [19.1-27.3] | 360 | 41.3 [36.5-46.3] | 258 | 34 [29.4-38.9] | 849 | 100 | -3.3 | -6.8[b] | 9.7[a] |
| Income | | | | | | | | | | | | | | | | | |
| Less than $25,000 | 135 | 27.6 [21.8-34.4] | 193 | 43.3 [36.4-50.5] | 96 | 25.6 [19.8-32.5] | 138 | 25.5 [20.1-31.9] | 163 | 40.3 [33.4-47.5] | 125 | 31.5 [25.2-38.5] | 431 | 100 | -2.1 | -3.0 | 5.9 |
| $25,000-$59,999 | 284 | 28 [23.8-32.6] | 367 | 53.3 [47.9-58.7] | 89 | 18.5 [14.3-23.6] | 265 | 26.2 [22.1-30.7] | 341 | 49.3 [43.9-54.8] | 133 | 23.6 [19.2-28.8] | 744 | 100 | -1.8 | -4.0 | 5.2 |
| $60,000-$99,999 | 353 | 41.7 [36.3-47.3] | 334 | 46.4 [40.8-52.0] | 64 | 11.2 [8.0-15.5] | 345 | 40.9 [35.5-46.5] | 306 | 39.1 [33.9-44.7] | 102 | 19.6 [15.3-24.8] | 757 | 100 | -0.8 | -7.2[b] | 8.3[a] |
| $100,000 or more | 519 | 51.1 [46.4-55.8] | 370 | 41.8 [37.2-46.6] | 44 | 6.4 [4.1-9.6] | 445 | 44.7 [40.1-49.4] | 412 | 44.7 [40.0-49.4] | 73 | 9.7 [7.0-13.3] | 938 | 100 | -6.4[b] | 2.8 | 3.4 |
| Age | | | | | | | | | | | | | | | | | |
| 18-29 | 67 | 33 [25.6-41.3] | 95 | 49.2 [40.8-57.6] | 36 | 17 [11.7-24.2] | 72 | 34.7 [27.1-43.2] | 80 | 43.7 [35.6-52.3] | 46 | 20.7 [14.9-28.2] | 200 | 100 | 1.8 | -5.5 | 3.7 |

| | | | | | | | | | | | | | | | | |
|---|---|---|---|---|---|---|---|---|---|---|---|---|---|---|---|---|
| 30-44 | 164 | 35.2 [29.8-41.0] | 260 | 47.6 [41.8-53.4] | 82 | 16.3 [12.3-21.3] | 158 | 30.9 [25.8-36.5] | 228 | 44.5 [38.8-50.3] | 119 | 23.1 [18.6-28.4] | 511 | 100 | -4.3 | -3.1 | 6.8[a] |
| 45-59 | 299 | 37.4 [32.6-42.3] | 339 | 48.4 [43.3-53.5] | 91 | 12.4 [9.6-16.0] | 261 | 33.2 [28.6-38.1] | 333 | 46.1 [41.1-51.2] | 138 | 19.7 [15.9-24.1] | 738 | 100 | -4.2 | -2.3 | 7.2[a] |
| 60+ | 761 | 51.7 [48.0-55.4] | 570 | 39.4 [35.8-43.1] | 84 | 8.5 [6.2-11.5] | 702 | 47 [43.3-50.7] | 581 | 41.1 [37.5-44.8] | 130 | 11.2 [8.7-14.2] | 1,421 | 100 | -4.7[b] | 1.7 | 2.7 |
| Household gun ownership | | | | | | | | | | | | | | | | | |
| No guns in home | 845 | 36.7 [33.6-39.8] | 897 | 47.9 [44.5-51.2] | 234 | 14.6 [12.3-17.2] | 805 | 34.6 [31.6-37.8] | 841 | 44.8 [41.5-48.1] | 331 | 19.7 [17.1-22.5] | 1,989 | 100 | -2.1 | -3.1 | 5.1[a] |
| Gun owner | 299 | 58.4 [51.8-64.8] | 207 | 35.3 [29.4-41.6] | 23 | 6.3 [3.4-11.4] | 247 | 43.3 [36.8-50.1] | 232 | 43.7 [37.2-50.4] | 47 | 12.5 [8.3-18.5] | 529 | 100 | -15.1[a] | 8.4[b] | 6.3[b] |
| Non-owner living with owner | 96 | 40.4 [31.4-50.2] | 99 | 43.9 [34.6-53.7] | 21 | 15.2 [8.8-25.0] | 96 | 44.8 [35.4-54.7] | 90 | 36.7 [28.0-46.3] | 32 | 18.3 [11.5-27.8] | 219 | 100 | 4.4 | -7.3 | 3.1 |
| Acquired firearms due to the coronavirus pandemic | | | | | | | | | | | | | | | | | |
| Non-gun owner | 992 | 36.6 [33.8-39.5] | 1057 | 47.7 [44.7-50.8] | 270 | 14.5 [12.4-16.9] | 946 | 35.3 [32.5-38.2] | 990 | 43.9 [40.9-47.0] | 386 | 19.6 [17.2-22.2] | 2,341 | 100 | -1.3 | -3.8[b] | 5.1[a] |
| Gun owner, no purchase | 294 | 58.5 [51.7-64.9] | 203 | 35.3 [29.3-41.8] | 22 | 6.2 [3.3-11.5] | 244 | 43.2 [36.6-50.1] | 227 | 43.8 [37.1-50.6] | 45 | 12.6 [8.3-18.7] | 519 | 100 | -15.2[a] | 8.5[b] | 6.3[b] |
| Gun owner, purchase | 5 | 58.3 [24.4-85.9] | 4 | 34.4 [10.5-70.2] | 1 | 7.2 [0.9-39.3] | 3 | 47.2 [16.1-80.6] | 5 | 41.1 [13.7-75.4] | 2 | 11.7 [2.5-40.9] | 10 | 100 | -11.1 | 6.6 | 4.5 |

See footnotes at the end of eAppendix 17.

eAppendix 11. Percentage of respondents worried about robbery happening to them before and during the coronavirus pandemic, California Safety and Wellbeing Survey 2020

| | Before the pandemic | | | | | | During the pandemic | | | | | | Total | | During - Before | | |
|---|---|---|---|---|---|---|---|---|---|---|---|---|---|---|---|---|---|
| | Not worried | | Somewhat worried | | Very worried | | Not worried | | Somewhat worried | | Very worried | | | | Not | Somewhat | Very |
| | n | % [95% CI] | n | % [95% CI] | n | % [95% CI] | n | % [95% CI] | n | % [95% CI] | n | % [95% CI] | n | % | PD | PD | PD |
| Total | 1048 | 33.5 [31.0-36.2] | 1469 | 50.2 [47.4-52.9] | 330 | 15.3 [13.3-17.6] | 997 | 30.8 [28.3-33.4] | 1364 | 46.3 [43.5-49.0] | 487 | 22 [19.6-24.5] | 2870 | 100 | -2.7 | -3.9[b] | 6.6[a] |
| Education | | | | | | | | | | | | | | | | | |
| Less than high school | 30 | 19.3 [13.2-27.4] | 67 | 44.9 [36.3-53.9] | 58 | 34 [26.1-42.8] | 27 | 18 [12.1-25.9] | 68 | 42.8 [34.3-51.8] | 62 | 37.9 [29.7-46.8] | 158 | 100 | -1.4 | -2.1 | 3.9 |
| High school | 97 | 30 [24.0-36.9] | 182 | 49.9 [43.1-56.7] | 79 | 18.5 [14.0-24.1] | 86 | 25.8 [20.1-32.5] | 157 | 42.9 [36.3-49.7] | 115 | 29.8 [24.0-36.3] | 364 | 100 | -4.2 | -7.0 | 11.2[a] |
| Some college | 362 | 40.4 [36.0-44.9] | 473 | 48.4 [44.0-52.9] | 96 | 10.3 [7.9-13.2] | 335 | 35.6 [31.4-40.1] | 445 | 46.3 [41.9-50.8] | 150 | 17.3 [14.1-21.0] | 938 | 100 | -4.7 | -2.1 | 7.0[a] |
| Bachelor's degree | 318 | 36.5 [31.5-41.8] | 389 | 51.1 [45.6-56.6] | 66 | 11.8 [8.0-17.1] | 298 | 34.2 [29.4-39.5] | 378 | 47.8 [42.3-53.3] | 97 | 17.4 [13.1-22.8] | 775 | 100 | -2.3 | -3.3 | 5.6[b] |
| Advanced degree | 241 | 34.4 [28.8-40.5] | 358 | 59.1 [52.8-65.1] | 31 | 6.1 [3.7-9.8] | 251 | 36.8 [31.0-43.0] | 316 | 53 [46.6-59.3] | 63 | 9.5 [6.7-13.4] | 635 | 100 | 2.3 | -6.1 | 3.4 |
| Gender | | | | | | | | | | | | | | | | | |
| Male | 624 | 38.7 [34.8-42.7] | 723 | 46.9 [42.8-50.9] | 148 | 13.4 [10.8-16.6] | 580 | 34.8 [31.0-38.7] | 691 | 45.1 [41.1-49.2] | 223 | 18.9 [15.8-22.5] | 1,506 | 100 | -3.9 | -1.7 | 5.5[a] |
| Female | 424 | 28.8 [25.6-32.3] | 746 | 53.2 [49.3-56.9] | 182 | 17 [14.2-20.3] | 417 | 27.2 [24.1-30.6] | 673 | 47.3 [43.5-51.1] | 264 | 24.7 [21.4-28.4] | 1,364 | 100 | -1.6 | -5.9[a] | 7.7[a] |
| Race/ethnicity | | | | | | | | | | | | | | | | | |
| White, Non-Hispanic | 697 | 44.5 [40.8-48.2] | 825 | 49.3 [45.6-53.1] | 81 | 5.3 [3.9-7.2] | 665 | 40.6 [37.0-44.3] | 793 | 48.4 [44.7-52.2] | 145 | 10 [7.9-12.5] | 1,615 | 100 | -3.9 | -0.9 | 4.7[a] |
| Black, Non-Hispanic | 44 | 32.6 [22.3-44.8] | 74 | 59.1 [46.8-70.3] | 9 | 8.3 [3.5-18.4] | 48 | 32.8 [22.6-45.0] | 68 | 56.2 [44.1-67.7] | 11 | 11 [5.3-21.3] | 127 | 100 | 0.2 | -2.9 | 2.6 |
| Asian, Non-Hispanic | 52 | 23.1 [16.9-30.8] | 118 | 59.7 [51.2-67.6] | 36 | 16.4 [11.0-23.8] | 53 | 23.3 [17.0-31.1] | 104 | 54.1 [45.6-62.4] | 51 | 22.5 [16.3-30.3] | 208 | 100 | 0.2 | -5.5 | 6.1 |
| Other & 2+ races, Non-Hispanic | 37 | 51.9 [35.0-68.5] | 29 | 40.1 [24.6-58.0] | 5 | 7.9 [2.8-20.2] | 36 | 45.4 [29.4-62.3] | 25 | 31.3 [17.9-48.8] | 10 | 23.3 [10.8-43.4] | 71 | 100 | -6.6 | -8.9 | 15.4[b] |
| Hispanic | 218 | 23.1 [19.1-27.5] | 423 | 46.6 [41.7-51.6] | 199 | 28.8 [24.4-33.6] | 195 | 20.4 [16.7-24.7] | 374 | 40 [35.3-45.0] | 270 | 37.9 [33.2-43.0] | 849 | 100 | -2.6 | -6.6[b] | 9.1[a] |
| Income | | | | | | | | | | | | | | | | | |
| Less than $25,000 | 119 | 25.1 [19.4-31.7] | 192 | 40.9 [34.1-48.0] | 110 | 30.1 [23.8-37.1] | 126 | 24.1 [18.7-30.6] | 168 | 37.8 [31.1-45.0] | 130 | 34.8 [28.3-41.9] | 431 | 100 | -0.9 | -3.0 | 4.7 |
| $25,000-$59,999 | 238 | 26 [21.7-30.7] | 409 | 55.2 [49.7-60.5] | 94 | 18.8 [14.6-23.7] | 212 | 21.9 [18.0-26.4] | 377 | 50.9 [45.4-56.3] | 150 | 26.8 [22.1-32.1] | 744 | 100 | -4.0 | -4.3 | 8.1[a] |
| $60,000-$99,999 | 291 | 33.8 [28.8-39.3] | 384 | 50.3 [44.7-55.9] | 79 | 15.4 [11.6-20.2] | 281 | 32.3 [27.3-37.7] | 358 | 44.5 [39.1-50.1] | 115 | 23.1 [18.4-28.5] | 757 | 100 | -1.5 | -5.8 | 7.6[a] |
| $100,000 or more | 400 | 41.4 [36.8-46.0] | 484 | 50.6 [45.9-55.3] | 47 | 7.3 [4.9-10.8] | 378 | 38.1 [33.7-42.7] | 461 | 47.7 [43.0-52.4] | 92 | 13.3 [10.1-17.3] | 938 | 100 | -3.3 | -2.9 | 5.9[a] |
| Age | | | | | | | | | | | | | | | | | |
| 18-29 | 76 | 40.4 [32.4-48.9] | 96 | 45.6 [37.4-54.0] | 26 | 13.3 [8.6-20.0] | 71 | 32.9 [25.6-41.1] | 80 | 42.4 [34.3-51.0] | 46 | 23.8 [17.4-31.8] | 200 | 100 | -7.5 | -3.1 | 10.6[a] |

| | | | | | | | | | | | | | | | | |
|---|---|---|---|---|---|---|---|---|---|---|---|---|---|---|---|---|
| 30-44 | 131 | 26.8 [21.9-32.3] | 273 | 51 [45.2-56.8] | 102 | 21.2 [16.7-26.5] | 124 | 26.1 [21.2-31.6] | 236 | 44.1 [38.4-49.9] | 145 | 28.6 [23.6-34.2] | 511 | 100 | -0.7 | -6.9[b] | 7.4[a] |
| 45-59 | 256 | 31.1 [26.8-35.8] | 369 | 51.4 [46.3-56.4] | 109 | 16.4 [12.9-20.5] | 233 | 28 [23.8-32.7] | 356 | 48.2 [43.1-53.3] | 144 | 22.8 [18.7-27.4] | 738 | 100 | -3.1 | -3.2 | 6.4[a] |
| 60+ | 585 | 39.1 [35.6-42.7] | 731 | 50.8 [47.0-54.5] | 93 | 9.1 [6.8-12.1] | 569 | 37.5 [34.0-41.1] | 692 | 49 [45.3-52.7] | 152 | 12.8 [10.3-15.9] | 1,421 | 100 | -1.6 | -1.8 | 3.8[b] |
| Household gun ownership | | | | | | | | | | | | | | | | | |
| No guns in home | 696 | 31.4 [28.4-34.5] | 1013 | 50.4 [47.1-53.8] | 267 | 17.4 [14.9-20.2] | 672 | 28.9 [26.0-31.9] | 939 | 46.9 [43.6-50.2] | 365 | 23.4 [20.6-26.4] | 1,989 | 100 | -2.5 | -3.5 | 6.0[a] |
| Gun owner | 232 | 45.2 [38.5-52.1] | 269 | 47.3 [40.6-54.0] | 28 | 7.5 [4.5-12.1] | 208 | 37.4 [31.0-44.2] | 261 | 48.1 [41.4-54.8] | 59 | 14.6 [10.1-20.5] | 529 | 100 | -7.9 | 0.8 | 7.1[a] |
| Non-owner living with owner | 72 | 31.4 [23.4-40.6] | 126 | 56.2 [46.5-65.5] | 20 | 12.2 [6.8-21.0] | 81 | 39.4 [30.3-49.2] | 100 | 38.8 [30.0-48.3] | 37 | 21.5 [13.9-31.7] | 219 | 100 | 8.0 | -17.4[a] | 9.3 |
| Acquired firearms due to the coronavirus pandemic | | | | | | | | | | | | | | | | | |
| Non-gun owner | 816 | 31.4 [28.7-34.3] | 1200 | 50.7 [47.6-53.7] | 302 | 16.7 [14.4-19.3] | 789 | 29.6 [27.0-32.4] | 1103 | 45.9 [42.9-49.0] | 428 | 23.3 [20.7-26.1] | 2,341 | 100 | -1.8 | -4.7[a] | 6.6[a] |
| Gun owner, no purchase | 228 | 45 [38.2-51.9] | 263 | 47.3 [40.6-54.2] | 28 | 7.7 [4.6-12.4] | 205 | 37 [30.6-43.9] | 256 | 48.4 [41.6-55.2] | 57 | 14.6 [10.1-20.7] | 519 | 100 | -8.0 | 1.0 | 6.9[a] |
| Gun owner, purchase | 4 | 55.6 [22.3-84.5] | 6 | 44.4 [15.5-77.7] | 0 | 0 [NA] | 3 | 52.7 [20.2-83.1] | 5 | 35.6 [11.2-70.8] | 2 | 11.7 [2.5-40.9] | 10 | 100 | -2.9 | -8.8 | 11.7 |

See footnotes at the end of eAppendix 17.

eAppendix 12. Percentage of respondents worried about police violence happening to them before and during the coronavirus pandemic, California Safety and Wellbeing Survey 2020

| | Before the pandemic | | | | | | During the pandemic | | | | | | Total | | During - Before | | |
|---|---|---|---|---|---|---|---|---|---|---|---|---|---|---|---|---|---|
| | Not worried | | Somewhat worried | | Very worried | | Not worried | | Somewhat worried | | Very worried | | | | Not | Somewhat | Very |
| | n | % [95% CI] | n | % [95% CI] | n | % [95% CI] | n | % [95% CI] | n | % [95% CI] | n | % [95% CI] | n | % | PD | PD | PD |
| Total | 1798 | 53.7 [50.8-56.5] | 737 | 30.5 [28.0-33.3] | 310 | 14.7 [12.7-17.0] | 1673 | 48.1 [45.4-50.9] | 739 | 30.9 [28.3-33.6] | 434 | 19.7 [17.5-22.2] | 2,870 | 100 | -5.5[a] | 0.3 | 5.0[a] |
| Education | | | | | | | | | | | | | | | | | |
| Less than high school | 56 | 33.4 [25.7-42.1] | 56 | 38.4 [30.1-47.5] | 41 | 25.6 [18.5-34.3] | 49 | 28.6 [21.3-37.1] | 58 | 39.8 [31.4-48.9] | 48 | 29.6 [22.1-38.4] | 158 | 100 | -4.9 | 1.4 | 4.0 |
| High school | 184 | 49.4 [42.6-56.2] | 107 | 31.2 [25.2-37.8] | 65 | 17.7 [12.9-23.9] | 156 | 39.5 [33.0-46.3] | 104 | 32.2 [26.2-39.0] | 97 | 26 [20.4-32.5] | 364 | 100 | -9.9[a] | 1.1 | 8.3[a] |
| Some college | 593 | 58.1 [53.6-62.6] | 242 | 28.2 [24.2-32.6] | 99 | 13 [10.0-16.6] | 577 | 55.7 [51.1-60.2] | 232 | 27.5 [23.5-31.8] | 122 | 16.1 [12.9-19.9] | 938 | 100 | -2.4 | -0.8 | 3.1 |
| Bachelor's degree | 519 | 58.1 [52.4-63.6] | 183 | 30 [24.9-35.6] | 71 | 11.4 [8.0-16.0] | 477 | 52.4 [46.9-58.0] | 199 | 28.7 [23.9-34.0] | 98 | 18.4 [14.0-23.7] | 775 | 100 | -5.7 | -1.3 | 7.0[a] |
| Advanced degree | 446 | 66.2 [59.6-72.2] | 149 | 27.1 [21.4-33.6] | 34 | 6.4 [3.8-10.5] | 414 | 59.4 [52.7-65.8] | 146 | 29.7 [23.5-36.8] | 69 | 9.9 [7.2-13.4] | 635 | 100 | -6.8 | 2.6 | 3.5 |
| Gender | | | | | | | | | | | | | | | | | |
| Male | 940 | 51.4 [47.3-55.5] | 390 | 31.2 [27.4-35.2] | 165 | 16.3 [13.2-20.0] | 898 | 47.8 [43.8-51.8] | 375 | 30.7 [26.9-34.7] | 219 | 20.2 [16.9-24.0] | 1,506 | 100 | -3.6 | -0.5 | 3.9 |
| Female | 858 | 55.7 [51.8-59.5] | 347 | 29.9 [26.5-33.7] | 145 | 13.2 [10.7-16.2] | 775 | 48.5 [44.7-52.3] | 364 | 31 [27.5-34.8] | 215 | 19.3 [16.4-22.6] | 1,364 | 100 | -7.2[a] | 1.1 | 6.1[a] |
| Race/ethnicity | | | | | | | | | | | | | | | | | |
| White, Non-Hispanic | 1227 | 72.6 [68.9-76.0] | 304 | 20.6 [17.6-24.0] | 75 | 6 [4.3-8.4] | 1166 | 67.4 [63.7-71.0] | 316 | 21.3 [18.3-24.7] | 124 | 10.4 [8.2-13.1] | 1,615 | 100 | -5.1[a] | 0.7 | 4.3[a] |
| Black, Non-Hispanic | 40 | 25.8 [17.1-36.9] | 46 | 37.9 [27.1-49.9] | 41 | 36.4 [25.3-49.1] | 40 | 28.8 [19.4-40.3] | 49 | 35.4 [25.1-47.3] | 38 | 35.8 [24.7-48.7] | 127 | 100 | 3.0 | -2.5 | -0.5 |
| Asian, Non-Hispanic | 107 | 50.6 [42.1-59.0] | 77 | 37.2 [29.3-45.9] | 24 | 12.2 [7.5-19.2] | 99 | 46.1 [37.8-54.6] | 74 | 37.5 [29.5-46.3] | 34 | 16.1 [10.8-23.3] | 208 | 100 | -4.5 | 0.3 | 3.9 |
| Other & 2+ races, Non-Hispanic | 48 | 73.1 [58.2-84.1] | 16 | 19 [10.0-33.1] | 6 | 5.6 [2.3-12.9] | 47 | 58 [40.2-74.0] | 16 | 35.3 [20.0-54.3] | 8 | 6.7 [3.0-14.3] | 71 | 100 | -15.0 | 16.2 | 1.1 |
| Hispanic | 376 | 34.9 [30.5-39.6] | 294 | 39.6 [34.8-44.6] | 164 | 23.5 [19.3-28.2] | 321 | 28 [24.0-32.4] | 284 | 38.5 [33.7-43.5] | 230 | 31.1 [26.6-36.0] | 849 | 100 | -6.9[a] | -1.1 | 7.6[a] |
| Income | | | | | | | | | | | | | | | | | |
| Less than $25,000 | 196 | 35.2 [29.0-41.9] | 129 | 36.6 [29.8-43.9] | 95 | 24 [18.2-31.0] | 188 | 34.1 [28.1-40.8] | 125 | 34.9 [28.2-42.1] | 110 | 27.7 [21.6-34.7] | 431 | 100 | -1.1 | -1.7 | 3.6 |
| $25,000-$59,999 | 435 | 47.8 [42.4-53.2] | 215 | 33.3 [28.4-38.7] | 90 | 18.7 [14.4-23.9] | 400 | 40 [35.0-45.2] | 209 | 33.8 [28.7-39.4] | 127 | 24.7 [20.1-30.0] | 744 | 100 | -7.8[a] | 0.5 | 6.0[b] |
| $60,000-$99,999 | 496 | 52.8 [47.1-58.4] | 189 | 34 [28.6-39.9] | 68 | 12.5 [9.0-17.0] | 452 | 46.2 [40.7-51.7] | 205 | 34.3 [28.9-40.1] | 99 | 19.5 [15.2-24.7] | 757 | 100 | -6.7[b] | 0.3 | 7.0[a] |
| $100,000 or more | 671 | 65 [60.2-69.5] | 204 | 24.4 [20.5-28.8] | 57 | 9.9 [7.0-13.7] | 633 | 59.8 [55.0-64.5] | 200 | 25.5 [21.4-30.0] | 98 | 13.7 [10.5-17.6] | 938 | 100 | -5.2 | 1.1 | 3.8 |
| Age | | | | | | | | | | | | | | | | | |
| 18-29 | 83 | 39.9 [32.0-48.4] | 78 | 41 [33.0-49.6] | 36 | 17.8 [12.2-25.2] | 69 | 34 [26.5-42.4] | 72 | 38.3 [30.3-46.9] | 57 | 26.9 [20.3-34.9] | 200 | 100 | -5.9 | -2.8 | 9.1[b] |

| | | | | | | | | | | | | | | | | |
|---|---|---|---|---|---|---|---|---|---|---|---|---|---|---|---|---|
| 30-44 | 248 | 46.8 [41.1-52.7] | 171 | 33.7 [28.5-39.4] | 83 | 18 [13.7-23.3] | 207 | 37.9 [32.5-43.6] | 177 | 36.3 [30.9-42.1] | 120 | 23.9 [19.2-29.3] | 511 | 100 | -8.9[a] | 2.6 | 5.9[b] |
| 45-59 | 437 | 53.1 [48.0-58.2] | 193 | 29.5 [25.0-34.5] | 103 | 16.1 [12.5-20.5] | 407 | 49.6 [44.6-54.7] | 181 | 27.6 [23.1-32.5] | 142 | 21.4 [17.5-26.0] | 738 | 100 | -3.5 | -1.9 | 5.3[b] |
| 60+ | 1030 | 69.8 [66.1-73.2] | 295 | 21.9 [18.9-25.2] | 88 | 7.9 [5.8-10.5] | 990 | 66.2 [62.5-69.8] | 309 | 23.8 [20.6-27.2] | 115 | 9.2 [7.1-11.9] | 1,421 | 100 | -3.6 | 1.9 | 1.4 |
| Household gun ownership | | | | | | | | | | | | | | | | | |
| No guns in home | 1162 | 48.7 [45.4-52.0] | 560 | 34.3 [31.1-37.6] | 248 | 15.9 [13.4-18.6] | 1079 | 43.8 [40.6-47.1] | 555 | 33.7 [30.5-37.1] | 555 | 21.4 [18.7-24.3] | 1,989 | 100 | -4.9[a] | -0.6 | 5.5[a] |
| Gun owner | 412 | 73.1 [66.3-79.0] | 92 | 18.3 [13.7-24.0] | 25 | 8.6 [4.8-15.0] | 390 | 65.6 [58.5-72.0] | 91 | 20.3 [15.3-26.5] | 91 | 13 [8.4-19.6] | 529 | 100 | -7.5 | 2.0 | 4.4 |
| Non-owner living with owner | 150 | 66.4 [56.6-75.0] | 53 | 19.7 [13.9-27.2] | 16 | 13.9 [7.5-24.3] | 129 | 54.5 [44.6-64.1] | 61 | 27.6 [19.7-37.3] | 61 | 17.9 [11.1-27.5] | 219 | 100 | -11.9[b] | 7.9 | 4.0 |
| Acquired firearms due to the coronavirus pandemic | | | | | | | | | | | | | | | | | |
| Non-gun owner | 1386 | 50.2 [47.1-53.2] | 645 | 32.7 [29.9-35.8] | 285 | 15.8 [13.5-18.3] | 1283 | 45 [42.0-48.0] | 648 | 32.8 [29.9-35.8] | 391 | 20.9 [18.5-23.6] | 2,341 | 100 | -5.2[a] | 0.0 | 5.1[a] |
| Gun owner, no purchase | 404 | 72.7 [65.7-78.7] | 90 | 18.5 [13.8-24.3] | 25 | 8.8 [4.9-15.3] | 382 | 64.9 [57.8-71.5] | 91 | 20.8 [15.7-27.1] | 41 | 13.1 [8.4-19.9] | 519 | 100 | -7.7 | 2.3 | 4.3 |
| Gun owner, purchase | 8 | 90.5 [65.6-98.0] | 2 | 9.5 [2.0-34.4] | 0 | 0 [NA] | 8 | 90.5 [65.6-98.0] | 0 | 0 [NA] | 2 | 9.5 [2.0-34.4] | 10 | 100 | 0.0 | -9.5 | 9.5 |

See footnotes at the end of eAppendix 17.

eAppendix 13. Percentage of respondents worried about an unintentional shooting happening to them before and during the coronavirus pandemic, California Safety and Wellbeing Survey 2020

| | Before the pandemic | | | | | | During the pandemic | | | | | | Total | | During - Before | | |
|---|---|---|---|---|---|---|---|---|---|---|---|---|---|---|---|---|---|
| | Not worried | | Somewhat worried | | Very worried | | Not worried | | Somewhat worried | | Very worried | | | | Not | Somewhat | Very |
| | n | % [95% CI] | n | % [95% CI] | n | % [95% CI] | n | % [95% CI] | n | % [95% CI] | n | % [95% CI] | n | % | PD | PD | PD |
| Total | 1837 | 56.2 [53.3-59.0] | 795 | 32.2 [29.6-34.9] | 216 | 10.5 [8.7-12.5] | 1697 | 51 [48.2-53.8] | 853 | 33.9 [31.2-36.6] | 298 | 14.2 [12.2-16.4] | 2,870 | 100 | -5.2[a] | 1.7 | 3.7[a] |
| Education | | | | | | | | | | | | | | | | | |
| Less than high school | 51 | 33 [25.2-41.8] | 62 | 40.9 [32.5-49.8] | 41 | 23.6 [16.8-32.0] | 46 | 27.5 [20.3-36.0] | 59 | 40.8 [32.3-49.9] | 50 | 30 [22.5-38.7] | 158 | 100 | -5.5 | -0.1 | 6.5 |
| High school | 186 | 50.8 [44.0-57.6] | 116 | 35.9 [29.5-42.9] | 56 | 11.2 [7.8-15.8] | 158 | 42.7 [36.1-49.5] | 128 | 38.4 [31.9-45.3] | 72 | 17.6 [13.1-23.3] | 364 | 100 | -8.1[b] | 2.5 | 6.4[b] |
| Some college | 627 | 65 [60.6-69.2] | 250 | 26.7 [23.0-30.9] | 54 | 7.5 [5.2-10.6] | 570 | 58.9 [54.5-63.3] | 275 | 29 [25.2-33.2] | 87 | 11.3 [8.6-14.6] | 938 | 100 | -6.1[b] | 2.3 | 3.8[b] |
| Bachelor's degree | 518 | 58.5 [52.7-64.0] | 210 | 32.7 [27.5-38.4] | 45 | 8.3 [5.3-12.9] | 490 | 55.3 [49.7-60.9] | 224 | 34 [28.8-39.7] | 59 | 10.1 [6.9-14.7] | 775 | 100 | -3.2 | 1.3 | 1.8 |
| Advanced degree | 455 | 66 [59.3-72.2] | 157 | 29.1 [23.2-35.9] | 20 | 4.6 [2.5-8.4] | 433 | 65 [58.3-71.2] | 167 | 30.8 [24.7-37.6] | 30 | 3.5 [2.2-5.6] | 635 | 100 | -1.0 | 1.7 | -1.1 |
| Gender | | | | | | | | | | | | | | | | | |
| Male | 1013 | 58.4 [54.2-62.4] | 383 | 31 [27.2-35.1] | 97 | 9.2 [6.9-12.2] | 938 | 54.5 [50.4-58.6] | 415 | 31.2 [27.4-35.2] | 143 | 13.3 [10.6-16.6] | 1,506 | 100 | -3.9 | 0.1 | 4.1[a] |
| Female | 824 | 54.2 [50.3-58.0] | 412 | 33.2 [29.7-36.9] | 119 | 11.6 [9.2-14.6] | 759 | 47.8 [44.0-51.5] | 438 | 36.3 [32.7-40.1] | 155 | 14.9 [12.3-18.1] | 1,364 | 100 | -6.4[a] | 3.1 | 3.3 |
| Race/ethnicity | | | | | | | | | | | | | | | | | |
| White, Non-Hispanic | 1236 | 76.8 [73.5-79.7] | 333 | 19.7 [16.9-22.7] | 40 | 2.8 [1.8-4.5] | 1138 | 67.6 [64.0-71.1] | 404 | 25.8 [22.7-29.2] | 64 | 5.6 [3.9-8.1] | 1,615 | 100 | -9.1[a] | 6.1[a] | 2.8[a] |
| Black, Non-Hispanic | 69 | 46.9 [35.3-58.8] | 47 | 42.3 [30.9-54.6] | 11 | 10.9 [5.0-22.2] | 70 | 46 [34.5-58.0] | 43 | 42.3 [30.8-54.7] | 14 | 11.7 [5.9-21.9] | 127 | 100 | -0.9 | 0.0 | 0.9 |
| Asian, Non-Hispanic | 96 | 42.8 [34.8-51.3] | 90 | 46.8 [38.4-55.4] | 21 | 10 [6.0-16.4] | 96 | 44.9 [36.7-53.4] | 82 | 42.5 [34.2-51.2] | 30 | 12.6 [8.1-19.0] | 208 | 100 | 2.0 | -4.3 | 2.6 |
| Other & 2+ races, Non-Hispanic | 46 | 60.3 [42.8-75.5] | 19 | 33.5 [19.2-51.6] | 6 | 6.2 [2.5-14.8] | 45 | 61.8 [44.2-76.7] | 20 | 31.6 [17.8-49.7] | 6 | 6.6 [2.2-18.1] | 71 | 100 | 1.5 | -1.9 | 0.4 |
| Hispanic | 390 | 38 [33.4-42.8] | 306 | 39.4 [34.6-44.4] | 138 | 20.2 [16.3-24.8] | 348 | 33.2 [28.9-37.9] | 304 | 38.9 [34.1-43.9] | 184 | 26.3 [22.0-31.0] | 849 | 100 | -4.8 | -0.5 | 6.0[b] |
| Income | | | | | | | | | | | | | | | | | |
| Less than $25,000 | 203 | 38 [31.6-44.9] | 135 | 37.3 [30.5-44.6] | 83 | 21 [15.7-27.7] | 190 | 33.7 [27.7-40.4] | 137 | 37.8 [30.9-45.1] | 95 | 25 [19.2-31.9] | 431 | 100 | -4.3 | 0.5 | 4.0 |
| $25,000-$59,999 | 442 | 48.2 [42.9-53.7] | 233 | 37.7 [32.5-43.2] | 64 | 13.2 [9.7-17.7] | 395 | 40.9 [35.9-46.1] | 251 | 41.7 [36.3-47.3] | 95 | 17.3 [13.6-21.8] | 744 | 100 | -7.4[b] | 4.0 | 4.1 |
| $60,000-$99,999 | 495 | 56.4 [50.7-62.0] | 215 | 33.1 [27.9-38.7] | 44 | 9.6 [6.4-14.0] | 474 | 53.3 [47.6-58.9] | 210 | 30.6 [25.7-36.0] | 69 | 15.7 [11.6-21.1] | 757 | 100 | -3.2 | -2.5 | 6.2[a] |
| $100,000 or more | 697 | 68 [63.2-72.5] | 212 | 26.2 [22.1-30.8] | 25 | 5.2 [3.1-8.5] | 638 | 62.6 [57.8-67.2] | 255 | 29.4 [25.2-34.0] | 39 | 7.1 [4.6-10.6] | 938 | 100 | -5.4 | 3.2 | 1.8 |
| Age | | | | | | | | | | | | | | | | | |
| 18-29 | 109 | 53.7 [45.2-61.9] | 62 | 32.5 [25.0-41.0] | 26 | 12 [7.7-18.2] | 102 | 49.9 [41.5-58.3] | 63 | 33.4 [25.9-41.8] | 33 | 15.9 [10.7-23.2] | 200 | 100 | -3.8 | 0.9 | 3.9 |

| | | | | | | | | | | | | | | | | | |
|---|---|---|---|---|---|---|---|---|---|---|---|---|---|---|---|---|---|
| 30-44 | 248 | 47 [41.3-52.8] | 183 | 36.1 [30.7-41.9] | 72 | 15.7 [11.7-20.7] | 222 | 40.6 [35.1-46.3] | 194 | 41.6 [35.9-47.5] | 88 | 16.6 [12.7-21.4] | 511 | 100 | -6.4 | 5.5 | 0.9 |
| 45-59 | 447 | 54.7 [49.5-59.7] | 223 | 35.1 [30.3-40.2] | 62 | 8.6 [6.1-12.1] | 412 | 48.6 [43.5-53.6] | 225 | 33.9 [29.2-38.9] | 96 | 16.5 [12.7-21.0] | 738 | 100 | -6.1[b] | -1.2 | 7.8[a] |
| 60+ | 1033 | 69.1 [65.3-72.6] | 327 | 24.9 [21.7-28.4] | 56 | 5.7 [3.9-8.2] | 961 | 65.3 [61.6-68.8] | 371 | 25.8 [22.7-29.2] | 81 | 8.3 [6.1-11.1] | 1,421 | 100 | -3.8 | 0.9 | 2.6 |
| Household gun ownership | | | | | | | | | | | | | | | | | |
| No guns in home | 1196 | 51.6 [48.3-55.0] | 601 | 35.5 [32.3-38.8] | 180 | 11.8 [9.7-14.2] | 1113 | 47.6 [44.3-50.9] | 612 | 35.2 [32.0-38.5] | 252 | 16.5 [14.1-19.3] | 1,989 | 100 | -4.1[b] | -0.3 | 4.7[a] |
| Gun owner | 417 | 75.5 [68.8-81.2] | 96 | 19.7 [14.7-25.9] | 14 | 4.4 [1.9-10.0] | 386 | 65.7 [58.6-72.1] | 119 | 26.3 [20.6-33.0] | 23 | 8 [4.4-14.2] | 529 | 100 | -9.9[a] | 6.7 | 3.5 |
| Non-owner living with owner | 144 | 60.7 [50.6-69.9] | 64 | 29 [21.1-38.6] | 11 | 10.3 [5.0-20.0] | 128 | 57.7 [47.8-67.0] | 76 | 29.6 [21.7-38.8] | 12 | 10.9 [5.6-20.2] | 219 | 100 | -3.0 | 0.5 | 0.7 |
| Acquired firearms due to the coronavirus pandemic | | | | | | | | | | | | | | | | | |
| Non-gun owner | 1420 | 52.7 [49.6-55.8] | 699 | 34.4 [31.5-37.4] | 202 | 11.6 [9.6-13.8] | 1311 | 48.4 [45.3-51.4] | 734 | 35.2 [32.3-38.3] | 275 | 15.3 [13.1-17.7] | 2,341 | 100 | -4.3[a] | 0.8 | 3.7[a] |
| Gun owner, no purchase | 408 | 75 [68.2-80.9] | 95 | 20 [14.9-26.4] | 14 | 4.6 [2.0-10.2] | 380 | 65.7 [58.4-72.2] | 115 | 26.1 [20.3-32.9] | 23 | 8.2 [4.5-14.5] | 519 | 100 | -9.4[b] | 6.1 | 3.6 |
| Gun owner, purchase | 9 | 95.5 [72.0-99.4] | 1 | 4.5 [0.6-28.0] | 0 | 0 [NA] | 6 | 65.6 [29.8-89.5] | 4 | 34.4 [10.5-70.2] | 0 | 0 [NA] | 10 | 100 | -29.9[b] | 29.9[b] | 0.0 |

See footnotes at the end of eAppendix 17.

eAppendix 14. Percentage of respondents worried about being hit by a stray bullet before and during the coronavirus pandemic, California Safety and Wellbeing Survey 2020

| | Before the pandemic | | | | | | During the pandemic | | | | | | Total | | During - Before | | |
|---|---|---|---|---|---|---|---|---|---|---|---|---|---|---|---|---|
| | Not worried | | Somewhat worried | | Very worried | | Not worried | | Somewhat worried | | Very worried | | | | Not | Somewhat | Very |
| | n | % [95% CI] | n | % [95% CI] | n | % [95% CI] | n | % [95% CI] | n | % [95% CI] | n | % [95% CI] | n | % | PD | PD | PD |
| Total | 1820 | 54.7 [51.9-57.5] | 799 | 33.7 [31.0-36.5] | 229 | 10.8 [9.1-12.7] | 1642 | 48.8 [46.0-51.6] | 898 | 35.8 [33.1-38.5] | 303 | 14.3 [12.3-16.5] | 2,870 | 100 | -5.9[a] | 2.0 | 3.5[a] |
| Education | | | | | | | | | | | | | | | | | |
| Less than high school | 37 | 21.4 [15.1-29.4] | 73 | 51.3 [42.5-60.1] | 47 | 25.9 [19.0-34.4] | 37 | 22 [15.7-30.1] | 64 | 42.7 [34.2-51.7] | 53 | 32.3 [24.5-41.2] | 158 | 100 | 0.7 | -8.6 | 6.3 |
| High school | 174 | 46.9 [40.2-53.8] | 123 | 39.9 [33.3-46.9] | 60 | 11.7 [8.4-15.9] | 143 | 39.2 [32.7-46.0] | 136 | 40.5 [34.0-47.4] | 78 | 18.8 [14.1-24.5] | 364 | 100 | -7.7 | 0.6 | 7.1[a] |
| Some college | 602 | 62.1 [57.7-66.4] | 266 | 28.8 [24.9-33.0] | 66 | 8.5 [6.2-11.6] | 531 | 54.7 [50.2-59.1] | 318 | 34.2 [30.2-38.5] | 84 | 10.5 [7.9-13.9] | 938 | 100 | -7.4[a] | 5.4[b] | 2.0 |
| Bachelor's degree | 553 | 66.9 [61.3-72.1] | 185 | 25.6 [21.0-30.8] | 36 | 7 [4.1-11.5] | 501 | 58.2 [52.5-63.7] | 216 | 33.4 [28.3-39.0] | 53 | 7.7 [4.9-12.0] | 775 | 100 | -8.7[a] | 7.8[a] | 0.8 |
| Advanced degree | 454 | 70.1 [63.5-75.9] | 152 | 26.8 [21.1-33.5] | 20 | 2.7 [1.5-4.6] | 430 | 66.7 [60.2-72.7] | 164 | 27.7 [22.0-34.3] | 35 | 4.7 [2.9-7.5] | 635 | 100 | -3.3 | 0.9 | 2.1 |
| Gender | | | | | | | | | | | | | | | | | |
| Male | 1003 | 56.5 [52.3-60.6] | 384 | 32.6 [28.7-36.7] | 108 | 10.1 [7.7-13.1] | 914 | 51 [46.9-55.1] | 430 | 34.6 [30.7-38.7] | 147 | 13.3 [10.6-16.6] | 1,506 | 100 | -5.5[b] | 2.0 | 3.2 |
| Female | 817 | 53 [49.2-56.9] | 415 | 34.8 [31.2-38.6] | 121 | 11.4 [9.1-14.2] | 728 | 46.8 [43.0-50.6] | 468 | 36.8 [33.3-40.6] | 156 | 15.1 [12.3-18.4] | 1,364 | 100 | -6.3[a] | 2.1 | 3.8[b] |
| Race/ethnicity | | | | | | | | | | | | | | | | | |
| White, Non-Hispanic | 1238 | 75.5 [72.1-78.7] | 326 | 20.8 [17.9-24.1] | 38 | 2.8 [1.7-4.5] | 1125 | 67.4 [63.8-70.8] | 411 | 27.6 [24.3-31.1] | 70 | 4.3 [3.0-6.1] | 1,615 | 100 | -8.1[a] | 6.8[a] | 1.5 |
| Black, Non-Hispanic | 67 | 41.9 [31.0-53.7] | 47 | 47.7 [35.9-59.8] | 13 | 10.4 [5.0-20.2] | 61 | 37.9 [27.4-49.5] | 54 | 52.9 [41.0-64.5] | 11 | 9.1 [4.1-18.9] | 127 | 100 | -4.1 | 5.2 | -1.3 |
| Asian, Non-Hispanic | 115 | 52.3 [43.7-60.7] | 72 | 38.2 [30.2-46.9] | 21 | 9.5 [5.6-15.7] | 98 | 46.7 [38.3-55.2] | 85 | 41.5 [33.4-50.1] | 24 | 11.7 [7.1-18.5] | 208 | 100 | -5.6 | 3.3 | 2.1 |
| Other & 2+ races, Non-Hispanic | 47 | 65.5 [48.6-79.3] | 21 | 30.9 [17.9-47.8] | 3 | 3.6 [1.0-12.5] | 45 | 54.1 [36.9-70.5] | 20 | 27.5 [15.4-44.1] | 6 | 18.4 [7.2-39.7] | 71 | 100 | -11.4 | -3.4 | 14.8[b] |
| Hispanic | 353 | 31.7 [27.5-36.2] | 333 | 45.4 [40.5-50.5] | 154 | 21.6 [17.7-26.1] | 313 | 28.5 [24.4-32.9] | 328 | 41.2 [36.4-46.2] | 192 | 27.9 [23.5-32.7] | 849 | 100 | -3.2 | -4.2 | 6.2[b] |
| Income | | | | | | | | | | | | | | | | | |
| Less than $25,000 | 182 | 34 [27.9-40.8] | 154 | 41.7 [34.7-49.0] | 87 | 21.3 [16.1-27.6] | 176 | 33.9 [27.7-40.8] | 149 | 38.9 [32.1-46.1] | 96 | 23.9 [18.2-30.6] | 431 | 100 | -0.1 | -2.8 | 2.6 |
| $25,000-$59,999 | 436 | 45.9 [40.7-51.3] | 232 | 38.8 [33.4-44.4] | 73 | 15.2 [11.5-19.9] | 373 | 36.3 [31.5-41.3] | 270 | 43.4 [38.0-48.9] | 95 | 19.8 [15.4-25.1] | 744 | 100 | -9.7[a] | 4.6 | 4.6 |
| $60,000-$99,999 | 486 | 49.7 [44.1-55.3] | 219 | 40.6 [35.0-46.5] | 46 | 9.2 [6.3-13.2] | 446 | 46.1 [40.7-51.7] | 233 | 38.6 [33.1-44.3] | 71 | 14.1 [10.3-18.8] | 757 | 100 | -3.6 | -2.0 | 4.9[b] |
| $100,000 or more | 716 | 71.1 [66.4-75.4] | 194 | 23.5 [19.6-27.9] | 23 | 4.8 [2.8-8.1] | 647 | 63.9 [59.2-68.4] | 246 | 28.2 [24.1-32.7] | 41 | 7.2 [4.7-10.7] | 938 | 100 | -7.2[a] | 4.7 | 2.3 |
| Age | | | | | | | | | | | | | | | | | |
| 18-29 | 109 | 53.8 [45.4-62.1] | 65 | 35.1 [27.4-43.6] | 24 | 10.3 [6.5-16.0] | 104 | 50 [41.6-58.4] | 64 | 33.2 [25.8-41.6] | 30 | 16 [10.5-23.4] | 200 | 100 | -3.8 | -1.9 | 5.6 |

| | | | | | | | | | | | | | | | | | |
|---|---|---|---|---|---|---|---|---|---|---|---|---|---|---|---|---|---|
| 30-44 | 244 | 46.3 [40.5-52.1] | 191 | 38.4 [32.9-44.2] | 72 | 14.5 [10.8-19.3] | 209 | 39.9 [34.3-45.7] | 197 | 40.1 [34.6-46.0] | 98 | 18.7 [14.5-23.7] | 511 | 100 | -6.4 | 1.8 | 4.2 |
| 45-59 | 445 | 51.5 [46.3-56.5] | 214 | 35.8 [30.8-41.0] | 72 | 11.5 [8.5-15.4] | 391 | 45.1 [40.2-50.1] | 245 | 37.5 [32.6-42.6] | 91 | 15.2 [11.6-19.6] | 738 | 100 | -6.3[b] | 1.7 | 3.7 |
| 60+ | 1022 | 67.5 [63.7-71.2] | 329 | 25.9 [22.5-29.6] | 61 | 6.2 [4.4-8.8] | 938 | 61.3 [57.5-65.0] | 392 | 30.9 [27.4-34.6] | 84 | 7.6 [5.6-10.3] | 1,421 | 100 | -6.2[a] | 5.0[b] | 1.3 |
| Household gun ownership | | | | | | | | | | | | | | | | | |
| No guns in home | 1184 | 49.9 [46.6-53.2] | 600 | 37 [33.7-40.3] | 191 | 12.4 [10.3-14.9] | 1080 | 45.2 [41.9-48.5] | 642 | 37.1 [33.9-40.4] | 249 | 16.5 [14.0-19.3] | 1,989 | 100 | -4.7[a] | 0.2 | 4.1[a] |
| Gun owner | 409 | 72.1 [65.2-78.0] | 107 | 23.6 [18.2-30.1] | 13 | 4.4 [1.9-9.9] | 362 | 62.9 [55.9-69.3] | 144 | 31.5 [25.4-38.3] | 22 | 5.6 [3.0-10.3] | 529 | 100 | -9.2[b] | 7.9[b] | 1.2 |
| Non-owner living with owner | 143 | 62.3 [52.3-71.3] | 62 | 28 [20.1-37.4] | 12 | 9.4 [4.6-18.5] | 130 | 55.2 [45.2-64.7] | 68 | 28.8 [21.0-38.0] | 20 | 15.8 [8.8-26.7] | 219 | 100 | -7.1 | 0.8 | 6.4 |
| Acquired firearms due to the coronavirus pandemic | | | | | | | | | | | | | | | | | |
| Non-gun owner | 1411 | 51.6 [48.5-54.6] | 692 | 35.5 [32.6-38.6] | 216 | 11.9 [10.0-14.1] | 1280 | 46.3 [43.3-49.3] | 754 | 36.5 [33.6-39.6] | 281 | 15.8 [13.6-18.4] | 2,341 | 100 | -5.3[a] | 1.0 | 3.9[a] |
| Gun owner, no purchase | 402 | 71.9 [64.8-78.0] | 104 | 23.7 [18.2-30.3] | 13 | 4.5 [1.9-10.1] | 356 | 62.8 [55.8-69.4] | 140 | 31.4 [25.2-38.4] | 22 | 5.7 [3.0-10.5] | 519 | 100 | -9.0[b] | 7.8[b] | 1.3 |
| Gun owner, purchase | 7 | 80.1 [47.0-94.8] | 3 | 19.9 [5.2-53.0] | 0 | 0 [NA] | 6 | 65.6 [29.8-89.5] | 4 | 34.4 [10.5-70.2] | 0 | 0 [NA] | 10 | 100 | -14.5 | 14.5 | 0.0 |

See footnotes at the end of eAppendix 17.

eAppendix 15. Percentage of respondents worried about a violent event happening to them in their home before and during the coronavirus pandemic, California Safety and Wellbeing Survey 2020

| | Before the pandemic | | | | | | During the pandemic | | | | | | Total | | During - Before | | |
|---|---|---|---|---|---|---|---|---|---|---|---|---|---|---|---|---|---|
| | Not worried | | Somewhat worried | | Very worried | | Not worried | | Somewhat worried | | Very worried | | | | Not | Somewhat | Very |
| | n | % [95% CI] | n | % [95% CI] | n | % [95% CI] | n | % [95% CI] | n | % [95% CI] | n | % [95% CI] | n | % | PD | PD | PD |
| Total | 2153 | 70.6 [67.9-73.1] | 583 | 22.5 [20.2-24.9] | 101 | 5.4 [4.1-7.1] | 2085 | 69.4 [66.7-72.0] | 615 | 21.9 [19.7-24.3] | 138 | 7.2 [5.7-8.9] | 2,870 | 100 | -1.2 | -0.6 | 1.7 |
| Education | | | | | | | | | | | | | | | | | |
| Less than high school | 86 | 54 [45.0-62.7] | 41 | 27.5 [20.3-36.1] | 26 | 15.7 [10.1-23.6] | 97 | 60.7 [51.7-69.1] | 31 | 19.3 [13.3-27.2] | 26 | 17.7 [11.7-25.8] | 158 | 100 | 6.7 | -8.2 | 2.0 |
| High school | 253 | 69 [62.2-75.0] | 81 | 23 [17.7-29.3] | 23 | 6 [3.5-10.2] | 231 | 62.9 [56.1-69.3] | 86 | 25.4 [19.9-31.9] | 36 | 8.7 [5.6-13.4] | 364 | 100 | -6.0 | 2.5 | 2.7 |
| Some college | 688 | 71.3 [67.0-75.2] | 210 | 24.3 [20.6-28.5] | 32 | 3.6 [2.3-5.7] | 676 | 70.7 [66.4-74.7] | 214 | 23.9 [20.2-28.1] | 41 | 4.5 [2.9-6.8] | 938 | 100 | -0.6 | -0.4 | 0.8 |
| Bachelor's degree | 615 | 77.8 [72.8-82.0] | 137 | 18.1 [14.4-22.4] | 17 | 2.8 [1.2-6.5] | 582 | 74.9 [69.8-79.3] | 164 | 19.3 [15.6-23.8] | 26 | 5.2 [2.9-9.3] | 775 | 100 | -2.9 | 1.3 | 2.5 |
| Advanced degree | 511 | 80.5 [75.1-84.9] | 114 | 17.4 [13.2-22.7] | 3 | 0.6 [0.2-2.5] | 499 | 78.7 [73.1-83.3] | 120 | 18.2 [13.8-23.7] | 9 | 1.9 [0.9-4.1] | 635 | 100 | -1.8 | 0.8 | 1.3 |
| Gender | | | | | | | | | | | | | | | | | |
| Male | 1169 | 71.8 [67.7-75.5] | 276 | 21.8 [18.5-25.5] | 43 | 4.8 [3.1-7.4] | 1115 | 69.7 [65.7-73.4] | 308 | 21.2 [18.0-24.8] | 68 | 7.5 [5.4-10.5] | 1,506 | 100 | -2.0 | -0.6 | 2.7 |
| Female | 984 | 69.5 [65.8-72.9] | 307 | 23.1 [20.1-26.4] | 58 | 6 [4.3-8.3] | 970 | 69.1 [65.5-72.6] | 307 | 22.5 [19.5-25.9] | 70 | 6.8 [5.0-9.2] | 1,364 | 100 | -0.4 | -0.6 | 0.8 |
| Race/ethnicity | | | | | | | | | | | | | | | | | |
| White, Non-Hispanic | 1305 | 77.8 [74.3-80.8] | 280 | 19 [16.1-22.1] | 22 | 2.5 [1.4-4.5] | 1231 | 72.5 [68.9-75.9] | 339 | 23 [19.9-26.4] | 34 | 3.5 [2.2-5.6] | 1,615 | 100 | -5.3[a] | 4.0[b] | 1.0 |
| Black, Non-Hispanic | 97 | 72.6 [60.5-82.1] | 26 | 25.1 [15.9-37.4] | 1 | 0.8 [0.1-5.7] | 99 | 77.7 [66.9-85.7] | 25 | 20.5 [12.8-31.3] | 1 | 0.6 [0.1-4.2] | 127 | 100 | 5.1 | -4.6 | -0.2 |
| Asian, Non-Hispanic | 146 | 69.8 [61.7-76.9] | 56 | 25.9 [19.4-33.7] | 5 | 3.4 [1.3-9.0] | 146 | 71.2 [63.0-78.2] | 52 | 23.8 [17.4-31.8] | 10 | 5 [2.4-10.0] | 208 | 100 | 1.3 | -2.0 | 1.5 |
| Other & 2+ races, Non-Hispanic | 55 | 84 [70.9-91.9] | 14 | 14.4 [7.0-27.4] | 0 | 0 [NA] | 54 | 72.8 [53.8-86.0] | 16 | 27 [13.8-46.1] | 1 | 0.2 [0.0-1.2] | 71 | 100 | -11.2 | 12.7 | 0.2 |
| Hispanic | 550 | 60.6 [55.6-65.4] | 207 | 25.6 [21.5-30.3] | 73 | 11 [8.1-14.9] | 555 | 63.3 [58.3-68.0] | 183 | 19.5 [16.0-23.7] | 92 | 14.2 [10.8-18.4] | 849 | 100 | 2.6 | -6.1[a] | 3.2 |
| Income | | | | | | | | | | | | | | | | | |
| Less than $25,000 | 274 | 57.7 [50.4-64.6] | 109 | 27 [21.3-33.6] | 35 | 10.9 [6.8-17.2] | 272 | 58.7 [51.5-65.6] | 103 | 23.3 [18.2-29.3] | 44 | 13.8 [9.0-20.4] | 431 | 100 | 1.1 | -3.7 | 2.9 |
| $25,000-$59,999 | 532 | 64.7 [59.2-69.9] | 166 | 25.3 [20.7-30.5] | 39 | 8.8 [5.9-12.8] | 525 | 65.6 [60.2-70.7] | 164 | 23.2 [18.8-28.2] | 46 | 9.4 [6.5-13.3] | 744 | 100 | 0.9 | -2.1 | 0.6 |
| $60,000-$99,999 | 586 | 73.4 [68.1-78.2] | 153 | 23.2 [18.7-28.4] | 15 | 2.8 [1.3-5.6] | 561 | 73.1 [67.9-77.8] | 169 | 22.3 [18.0-27.2] | 23 | 4.4 [2.5-7.5] | 757 | 100 | -0.3 | -0.9 | 1.6 |
| $100,000 or more | 761 | 77.6 [73.2-81.4] | 155 | 18.5 [15.1-22.6] | 12 | 2.7 [1.3-5.6] | 727 | 73.8 [69.1-77.9] | 179 | 20.4 [16.7-24.6] | 25 | 4.8 [2.9-8.0] | 938 | 100 | -3.8 | 1.8 | 2.1 |
| Age | | | | | | | | | | | | | | | | | |
| 18-29 | 142 | 72.4 [64.5-79.1] | 46 | 21 [15.1-28.5] | 9 | 5.1 [2.5-10.1] | 140 | 69 [60.7-76.2] | 46 | 24 [17.5-32.0] | 11 | 5.6 [2.8-10.9] | 200 | 100 | -3.4 | 3.0 | 0.5 |

| | | | | | | | | | | | | | | | | | |
|---|---|---|---|---|---|---|---|---|---|---|---|---|---|---|---|---|---|
| 30-44 | 337 | 64.5 [58.7-69.9] | 134 | 26.4 [21.6-31.8] | 29 | 6.8 [4.2-10.9] | 324 | 63 [57.2-68.5] | 132 | 23.9 [19.4-29.1] | 44 | 10.9 [7.6-15.5] | 511 | 100 | -1.5 | -2.5 | 4.1 |
| 45-59 | 537 | 69.4 [64.3-74.0] | 161 | 22.7 [18.7-27.3] | 32 | 6.7 [4.3-10.3] | 521 | 70.8 [65.9-75.2] | 163 | 20.4 [16.6-24.7] | 48 | 7.7 [5.3-11.0] | 738 | 100 | 1.4 | -2.4 | 1.0 |
| 60+ | 1137 | 77.2 [73.6-80.5] | 242 | 18.9 [16.0-22.2] | 31 | 2.8 [1.5-5.2] | 1100 | 75.3 [71.7-78.6] | 274 | 20 [17.1-23.3] | 35 | 3.4 [2.0-5.9] | 1,421 | 100 | -1.9 | 1.1 | 0.6 |
| Household gun ownership | | | | | | | | | | | | | | | | | |
| No guns in home | 1481 | 69.7 [66.4-72.7] | 401 | 22.9 [20.2-25.9] | 84 | 5.8 [4.3-7.8] | 1448 | 69.2 [66.0-72.3] | 411 | 21.8 [19.2-24.7] | 108 | 7.4 [5.7-9.4] | 1,989 | 100 | -0.4 | -1.1 | 1.5 |
| Gun owner | 429 | 78 [71.5-83.4] | 94 | 19.1 [14.2-25.2] | 3 | 2.4 [0.7-8.1] | 404 | 73.5 [66.7-79.4] | 114 | 20.4 [15.6-26.2] | 9 | 5.8 [2.6-12.5] | 529 | 100 | -4.5 | 1.3 | 3.4 |
| Non-owner living with owner | 158 | 71.2 [61.7-79.2] | 51 | 21.6 [15.0-30.0] | 10 | 7.2 [3.0-16.2] | 153 | 69 [59.1-77.4] | 52 | 22.9 [15.8-32.0] | 13 | 8 [3.7-16.7] | 219 | 100 | -2.3 | 1.4 | 0.8 |
| Acquired firearms due to the coronavirus pandemic | | | | | | | | | | | | | | | | | |
| Non-gun owner | 1724 | 69.2 [66.3-72.0] | 489 | 23.1 [20.6-25.8] | 98 | 6 [4.5-7.8] | 1681 | 68.7 [65.7-71.5] | 501 | 22.2 [19.7-24.8] | 129 | 7.4 [5.8-9.3] | 2,341 | 100 | -0.6 | -0.9 | 1.4 |
| Gun owner, no purchase | 419 | 77.5 [70.8-83.0] | 94 | 19.6 [14.6-25.8] | 3 | 2.5 [0.7-8.3] | 397 | 73.2 [66.3-79.2] | 111 | 20.5 [15.6-26.5] | 9 | 6 [2.7-12.8] | 519 | 100 | -4.3 | 0.9 | 3.5 |
| Gun owner, purchase | 10 | 100 [NA] | 0 | 0 [NA] | 0 | 0 [NA] | 7 | 84.6 [56.0-96.0] | 3 | 15.4 [4.0-44.0] | 0 | 0 [NA] | 10 | 100 | -15.4 | 15.4 | 0.0 |

See footnotes at the end of eAppendix 17.

eAppendix 16. Percentage of respondents worried about a violent event happening to them in their neighborhood before and during the coronavirus pandemic, California Safety and Wellbeing Survey 2020

| | Before the pandemic | | | | | | During the pandemic | | | | | | Total | | During - Before | | |
|---|---|---|---|---|---|---|---|---|---|---|---|---|---|---|---|---|---|
| | Not worried | | Somewhat worried | | Very worried | | Not worried | | Somewhat worried | | Very worried | | | | Not | Somewhat | Very |
| | n | % [95% CI] | n | % [95% CI] | n | % [95% CI] | n | % [95% CI] | n | % [95% CI] | n | % [95% CI] | n | % | PD | PD | PD |
| Total | 1617 | 50 [47.3-52.8] | 1042 | 39.6 [36.9-42.4] | 188 | 9.2 [7.6-11.2] | 1474 | 46.4 [43.7-49.2] | 1131 | 40.9 [38.2-43.7] | 237 | 11.3 [9.5-13.4] | 2,870 | 100 | -3.6[b] | 1.3 | 2.1 |
| Education | | | | | | | | | | | | | | | | | |
| Less than high school | 44 | 28.4 [21.0-37.1] | 71 | 46.4 [37.7-55.3] | 38 | 21.8 [15.3-30.1] | 53 | 32.3 [24.7-41.1] | 60 | 39 [30.8-48.0] | 39 | 24.8 [17.8-33.4] | 158 | 100 | 4.0 | -7.4 | 3.0 |
| High school | 158 | 45.4 [38.7-52.3] | 164 | 43.3 [36.7-50.2] | 39 | 10.4 [6.9-15.5] | 138 | 40.8 [34.2-47.7] | 165 | 42.5 [35.9-49.3] | 57 | 15.8 [11.3-21.6] | 364 | 100 | -4.6 | -0.8 | 5.4 |
| Some college | 511 | 52.8 [48.3-57.2] | 353 | 39.9 [35.5-44.4] | 67 | 6.7 [4.9-9.0] | 473 | 49.5 [45.0-53.9] | 385 | 41.8 [37.4-46.3] | 72 | 7.9 [5.8-10.8] | 938 | 100 | -3.3 | 1.9 | 1.3 |
| Bachelor's degree | 486 | 58.8 [53.1-64.2] | 253 | 34.9 [29.8-40.3] | 32 | 5.7 [3.2-10.1] | 427 | 51.4 [45.9-56.9] | 297 | 41 [35.6-46.6] | 45 | 6.2 [3.9-9.7] | 775 | 100 | -7.4[b] | 6.1 | 0.5 |
| Advanced degree | 418 | 63.4 [56.9-69.5] | 201 | 31.9 [26.2-38.3] | 12 | 4 [1.7-8.8] | 383 | 57 [50.4-63.4] | 224 | 38.2 [32.0-44.8] | 24 | 4.1 [2.1-7.7] | 635 | 100 | -6.4 | 6.3 | 0.1 |
| Gender | | | | | | | | | | | | | | | | | |
| Male | 891 | 51.8 [47.7-55.9] | 515 | 38.3 [34.3-42.4] | 87 | 8.8 [6.5-11.9] | 820 | 48.9 [44.9-53.0] | 550 | 38.1 [34.2-42.1] | 119 | 11.4 [8.8-14.6] | 1,506 | 100 | -2.8 | -0.2 | 2.5 |
| Female | 726 | 48.5 [44.7-52.3] | 527 | 40.8 [37.1-44.6] | 101 | 9.6 [7.4-12.3] | 654 | 44.2 [40.5-48.0] | 581 | 43.5 [39.7-47.3] | 118 | 11.3 [8.9-14.1] | 1,364 | 100 | -4.3 | 2.6 | 1.7 |
| Race/ethnicity | | | | | | | | | | | | | | | | | |
| White, Non-Hispanic | 1084 | 64 [60.2-67.6] | 480 | 31.9 [28.5-35.5] | 42 | 2.9 [1.8-4.5] | 975 | 57.3 [53.5-61.0] | 566 | 36.9 [33.3-40.6] | 64 | 4.5 [3.1-6.4] | 1,615 | 100 | -6.7[a] | 5.0[b] | 1.6 |
| Black, Non-Hispanic | 62 | 38.4 [27.9-50.1] | 57 | 54.1 [42.2-65.6] | 7 | 7.3 [2.8-17.7] | 67 | 45.5 [34.1-57.4] | 52 | 46.1 [34.4-58.2] | 7 | 8.2 [3.0-20.4] | 127 | 100 | 7.1 | -8.0 | 1.0 |
| Asian, Non-Hispanic | 103 | 48.9 [40.5-57.4] | 90 | 41.4 [33.3-50.0] | 15 | 9.7 [5.5-16.6] | 91 | 41.1 [33.0-49.6] | 100 | 51 [42.5-59.4] | 16 | 7.1 [4.0-12.5] | 208 | 100 | -7.9 | 9.5 | -2.5 |
| Other & 2+ races, Non-Hispanic | 48 | 63.6 [45.6-78.4] | 21 | 35.3 [20.6-53.5] | 1 | 0.8 [0.1-5.6] | 41 | 59.9 [42.4-75.1] | 28 | 39.2 [24.0-56.8] | 2 | 1 [0.2-5.0] | 71 | 100 | -3.7 | 3.9 | 0.2 |
| Hispanic | 320 | 34.3 [29.9-39.1] | 394 | 46.1 [41.2-51.1] | 123 | 17.9 [14.2-22.2] | 300 | 34.5 [29.9-39.3] | 385 | 40.8 [36.1-45.7] | 148 | 22.8 [18.6-27.6] | 849 | 100 | 0.1 | -5.3 | 5.0 |
| Income | | | | | | | | | | | | | | | | | |
| Less than $25,000 | 168 | 33.4 [27.2-40.3] | 188 | 44.5 [37.5-51.6] | 67 | 18.8 [13.5-25.5] | 166 | 36.2 [29.6-43.4] | 180 | 39 [32.4-46.0] | 76 | 21.2 [15.6-28.1] | 431 | 100 | 2.8 | -5.5 | 2.4 |
| $25,000-$59,999 | 359 | 39.8 [34.8-45.1] | 309 | 43.8 [38.5-49.3] | 69 | 15 [11.2-20.0] | 323 | 37.8 [32.8-43.1] | 339 | 44.5 [39.2-49.9] | 74 | 16.3 [12.2-21.4] | 744 | 100 | -2.0 | 0.7 | 1.2 |
| $60,000-$99,999 | 450 | 51.6 [46.0-57.2] | 277 | 43.8 [38.2-49.5] | 28 | 4.5 [2.8-7.3] | 417 | 48.3 [42.8-53.9] | 285 | 42.1 [36.6-47.8] | 51 | 8.9 [6.0-12.8] | 757 | 100 | -3.3 | -1.7 | 4.3[a] |
| $100,000 or more | 640 | 62 [57.1-66.6] | 268 | 32.6 [28.2-37.3] | 24 | 4.6 [2.7-7.8] | 568 | 54.7 [49.9-59.4] | 327 | 38.6 [34.1-43.4] | 36 | 5.8 [3.7-9.0] | 938 | 100 | -7.2[a] | 6.0[b] | 1.2 |
| Age | | | | | | | | | | | | | | | | | |
| 18-29 | 89 | 42.9 [34.9-51.4] | 89 | 47.2 [38.9-55.7] | 20 | 9 [5.4-14.7] | 87 | 43.6 [35.5-52.0] | 91 | 46.5 [38.2-55.0] | 19 | 8.4 [4.8-14.3] | 200 | 100 | 0.6 | -0.7 | -0.6 |

| | | | | | | | | | | | | | | | | | |
|---|---|---|---|---|---|---|---|---|---|---|---|---|---|---|---|---|---|
| 30-44 | 212 | 44.1 [38.4-50.0] | 228 | 40.6 [35.1-46.3] | 63 | 13.9 [10.1-18.8] | 200 | 40.2 [34.6-46.0] | 223 | 40.5 [35.0-46.3] | 81 | 18 [13.7-23.2] | 511 | 100 | -4.0 | -0.1 | 4.1 |
| 45-59 | 400 | 49.4 [44.3-54.4] | 279 | 41.5 [36.5-46.7] | 54 | 8.1 [5.6-11.6] | 347 | 44 [39.0-49.0] | 315 | 44.1 [39.0-49.2] | 72 | 11.1 [8.3-14.7] | 738 | 100 | -5.4 | 2.5 | 3.0 |
| 60+ | 916 | 61.3 [57.5-65.0] | 446 | 32.1 [28.7-35.7] | 51 | 5.4 [3.6-8.0] | 840 | 57.4 [53.6-61.1] | 502 | 34.9 [31.5-38.4] | 65 | 6 [4.0-8.9] | 1,421 | 100 | -4.0 | 2.7 | 0.6 |
| Household gun ownership | | | | | | | | | | | | | | | | | |
| No guns in home | 1058 | 47.1 [43.8-50.4] | 764 | 41.3 [38.0-44.6] | 154 | 10.6 [8.5-13.0] | 979 | 44.4 [41.1-47.7] | 799 | 41.1 [37.9-44.4] | 194 | 13.2 [10.9-15.8] | 1,989 | 100 | -2.8 | -0.2 | 2.6 |
| Gun owner | 371 | 65.8 [58.8-72.1] | 147 | 31.4 [25.2-38.2] | 8 | 2.6 [1.0-6.7] | 325 | 56.6 [49.7-63.3] | 185 | 37.7 [31.4-44.5] | 15 | 5.4 [2.5-11.6] | 529 | 100 | -9.2[b] | 6.4 | 2.9 |
| Non-owner living with owner | 126 | 54.1 [44.2-63.6] | 81 | 36.4 [27.6-46.2] | 12 | 9.5 [4.5-19.0] | 116 | 54.2 [44.4-63.6] | 90 | 38.9 [30.0-48.6] | 13 | 6.9 [3.0-15.3] | 219 | 100 | 0.1 | 2.5 | -2.6 |
| Acquired firearms due to the coronavirus pandemic | | | | | | | | | | | | | | | | | |
| Non-gun owner | 1246 | 47.2 [44.2-50.3] | 895 | 41.1 [38.1-44.1] | 180 | 10.4 [8.6-12.6] | 1149 | 44.6 [41.6-47.7] | 946 | 41.4 [38.5-44.5] | 222 | 12.4 [10.4-14.7] | 2,341 | 100 | -2.6 | 0.4 | 1.9 |
| Gun owner, no purchase | 365 | 65.8 [58.8-72.3] | 143 | 31.2 [25.0-38.2] | 8 | 2.6 [1.0-6.9] | 321 | 56.6 [49.6-63.4] | 179 | 37.6 [31.1-44.5] | 15 | 5.6 [2.5-11.9] | 519 | 100 | -9.2[b] | 6.4 | 2.9 |
| Gun owner, purchase | 6 | 62 [26.5-88.1] | 4 | 38 [11.9-73.5] | 0 | 0 [NA] | 4 | 54.7 [21.5-84.2] | 6 | 45.3 [15.8-78.5] | 0 | 0 [NA] | 10 | 100 | -7.3 | 7.3 | 0.0 |

See footnotes at the end of eAppendix 17.

eAppendix 17. Percentage of respondents worried about a violent event happening to them somewhere else before and during the coronavirus pandemic, California Safety and Wellbeing Survey 2020

| | Before the pandemic | | | | | | During the pandemic | | | | | | Total | | During - Before | | |
|---|---|---|---|---|---|---|---|---|---|---|---|---|---|---|---|---|---|
| | Not worried | | Somewhat worried | | Very worried | | Not worried | | Somewhat worried | | Very worried | | | | Not | Somewhat | Very |
| | n | % [95% CI] | n | % [95% CI] | n | % [95% CI] | n | % [95% CI] | n | % [95% CI] | n | % [95% CI] | n | % | PD | PD | PD |
| Total | 890 | 27.1 [24.8-29.7] | 1531 | 51.7 [48.9-54.4] | 419 | 19.7 [17.4-22.2] | 883 | 26.7 [24.3-29.1] | 1434 | 48.3 [45.5-51.0] | 522 | 23.5 [21.1-26.0] | 2,870 | 100 | -0.5 | -3.4[b] | 3.8[a] |
| Education | | | | | | | | | | | | | | | | | |
| Less than high school | 29 | 19.2 [12.9-27.4] | 64 | 41.9 [33.5-50.9] | 60 | 35.2 [27.2-44.0] | 32 | 20.8 [14.3-29.1] | 56 | 35.9 [27.9-44.8] | 62 | 38.8 [30.6-47.8] | 158 | 100 | 1.6 | -6.0 | 3.6 |
| High school | 102 | 29.2 [23.3-35.9] | 173 | 46.9 [40.2-53.8] | 82 | 22.4 [17.1-28.8] | 92 | 24.6 [19.2-31.0] | 157 | 45.4 [38.7-52.3] | 111 | 29 [23.2-35.6] | 364 | 100 | -4.6 | -1.6 | 6.6 |
| Some college | 283 | 27.1 [23.4-31.2] | 520 | 55.2 [50.7-59.6] | 127 | 16.8 [13.5-20.7] | 267 | 26.1 [22.4-30.1] | 503 | 52.1 [47.6-56.5] | 160 | 20.9 [17.2-25.0] | 938 | 100 | -1.0 | -3.1 | 4.0 |
| Bachelor's degree | 268 | 28.7 [24.2-33.8] | 415 | 56.5 [50.8-61.9] | 89 | 13.6 [9.7-18.8] | 264 | 26.9 [22.6-31.7] | 404 | 56.3 [50.7-61.7] | 104 | 15.7 [11.8-20.6] | 775 | 100 | -1.9 | -0.2 | 2.1 |
| Advanced degree | 208 | 31 [25.5-37.2] | 359 | 55 [48.5-61.3] | 61 | 12.8 [8.5-18.6] | 228 | 37.2 [31.2-43.7] | 314 | 47.2 [40.8-53.7] | 85 | 14.3 [10.3-19.6] | 635 | 100 | 6.2 | -7.8[b] | 1.6 |
| Gender | | | | | | | | | | | | | | | | | |
| Male | 507 | 29.7 [26.2-33.5] | 796 | 52.8 [48.7-56.8] | 185 | 15.9 [12.9-19.4] | 498 | 28.1 [24.7-31.8] | 751 | 48.6 [44.5-52.7] | 240 | 21.5 [18.1-25.4] | 1,506 | 100 | -1.6 | -4.2 | 5.6[a] |
| Female | 383 | 24.8 [21.7-28.2] | 735 | 50.6 [46.8-54.5] | 234 | 23.1 [19.8-26.7] | 385 | 25.3 [22.2-28.7] | 683 | 48 [44.2-51.8] | 282 | 25.3 [22.0-28.8] | 1,364 | 100 | 0.5 | -2.7 | 2.2 |
| Race/ethnicity | | | | | | | | | | | | | | | | | |
| White, Non-Hispanic | 592 | 34.7 [31.2-38.3] | 885 | 54.5 [50.7-58.2] | 129 | 9.6 [7.4-12.4] | 586 | 33.4 [30.1-37.0] | 841 | 52.5 [48.7-56.2] | 177 | 12.7 [10.3-15.6] | 1,615 | 100 | -1.3 | -2.0 | 3.1[b] |
| Black, Non-Hispanic | 40 | 24.6 [16.1-35.6] | 56 | 43.8 [32.4-55.9] | 29 | 30.5 [20.0-43.4] | 44 | 31.2 [21.3-43.2] | 62 | 47.7 [36.0-59.7] | 19 | 19.9 [11.3-32.7] | 127 | 100 | 6.6 | 3.9 | -10.6 |
| Asian, Non-Hispanic | 51 | 24.1 [17.6-32.1] | 118 | 53.2 [44.6-61.6] | 38 | 21.8 [15.3-30.2] | 47 | 22.6 [16.3-30.5] | 109 | 51.9 [43.4-60.3] | 51 | 24.6 [18.1-32.6] | 208 | 100 | -1.5 | -1.3 | 2.8 |
| Other & 2+ races, Non-Hispanic | 27 | 34.6 [20.6-52.0] | 38 | 56.9 [39.7-72.5] | 5 | 8.2 [2.7-22.4] | 28 | 34.5 [20.5-51.9] | 37 | 57.1 [40.0-72.7] | 6 | 8.3 [2.8-22.4] | 71 | 100 | -0.1 | 0.3 | 0.2 |
| Hispanic | 180 | 19 [15.4-23.3] | 434 | 48.5 [43.5-53.4] | 218 | 30.2 [25.7-35.0] | 178 | 18.7 [15.1-22.9] | 385 | 40.9 [36.2-45.8] | 269 | 38 [33.2-43.1] | 849 | 100 | -0.4 | -7.6[a] | 7.9[a] |
| Income | | | | | | | | | | | | | | | | | |
| Less than $25,000 | 111 | 22.8 [17.4-29.3] | 209 | 47.2 [40.2-54.4] | 102 | 26.4 [20.6-33.3] | 115 | 26.3 [20.3-33.3] | 191 | 37.4 [31.0-44.2] | 117 | 32.6 [26.0-39.8] | 431 | 100 | 3.5 | -9.8[a] | 6.1 |
| $25,000-$59,999 | 194 | 18.1 [14.8-22.0] | 409 | 52.7 [47.2-58.1] | 133 | 27.6 [22.6-33.3] | 172 | 17 [13.6-21.0] | 405 | 49.6 [44.1-55.0] | 156 | 31.5 [26.4-37.2] | 744 | 100 | -1.1 | -3.2 | 3.9 |
| $60,000-$99,999 | 261 | 30.5 [25.5-35.9] | 396 | 51.8 [46.2-57.4] | 96 | 16.6 [12.6-21.6] | 264 | 28.1 [23.5-33.2] | 353 | 49.7 [44.1-55.3] | 134 | 21.1 [16.8-26.1] | 757 | 100 | -2.4 | -2.1 | 4.5 |
| $100,000 or more | 324 | 32.5 [28.3-37.0] | 517 | 52.6 [47.9-57.3] | 88 | 13.8 [10.5-18.0] | 332 | 32 [27.8-36.4] | 485 | 50.9 [46.1-55.6] | 115 | 16.3 [12.8-20.5] | 938 | 100 | -0.5 | -1.8 | 2.5 |
| Age | | | | | | | | | | | | | | | | | |
| 18-29 | 34 | 16.6 [11.1-24.0] | 111 | 55 [46.5-63.2] | 51 | 26.4 [19.6-34.5] | 42 | 18.3 [12.8-25.5] | 102 | 52.6 [44.2-60.9] | 52 | 26.9 [20.0-35.1] | 200 | 100 | 1.7 | -2.4 | 0.5 |

| | | | | | | | | | | | | | | | | | |
|---|---|---|---|---|---|---|---|---|---|---|---|---|---|---|---|---|---|
| 30-44 | 122 | 26.4 [21.5-32.0] | 279 | 49.9 [44.1-55.7] | 101 | 22.2 [17.6-27.6] | 134 | 27.3 [22.3-32.9] | 243 | 44.6 [38.9-50.4] | 125 | 26.5 [21.6-32.1] | 511 | 100 | 0.9 | -5.3 | 4.3 |
| 45-59 | 216 | 27.6 [23.3-32.4] | 393 | 51.5 [46.4-56.6] | 122 | 19.7 [15.7-24.3] | 192 | 24.3 [20.3-28.9] | 376 | 48.2 [43.2-53.3] | 163 | 26 [21.6-30.9] | 738 | 100 | -3.3 | -3.2 | 6.3[b] |
| 60+ | 518 | 33.8 [30.5-37.2] | 748 | 51.7 [48.0-55.4] | 145 | 12.9 [10.2-16.1] | 515 | 33.2 [30.0-36.7] | 713 | 49.7 [46.0-53.4] | 182 | 15.6 [12.8-19.0] | 1,421 | 100 | -0.5 | -2.1 | 2.8 |
| Household gun ownership | | | | | | | | | | | | | | | | | |
| No guns in home | 576 | 24.9 [22.2-27.9] | 1064 | 51.5 [48.1-54.8] | 330 | 22.2 [19.4-25.3] | 594 | 26.2 [23.4-29.2] | 980 | 46.6 [43.3-49.9] | 394 | 25.5 [22.6-28.7] | 1,989 | 100 | 1.3 | -4.9[a] | 3.3 |
| Gun owner | 215 | 38.5 [32.1-45.2] | 282 | 54.2 [47.4-60.8] | 30 | 6.9 [4.4-10.6] | 193 | 29.1 [23.9-35.0] | 272 | 53.3 [46.5-59.9] | 62 | 17.2 [12.2-23.9] | 529 | 100 | -9.4[a] | -0.9 | 10.4[a] |
| Non-owner living with owner | 61 | 27.9 [19.9-37.7] | 122 | 49.8 [40.2-59.5] | 36 | 22.2 [14.6-32.3] | 65 | 30.2 [21.9-40.0] | 114 | 48.2 [38.6-57.9] | 40 | 21.6 [14.4-31.3] | 219 | 100 | 2.3 | -1.7 | -0.6 |
| Acquired firearms due to the coronavirus pandemic | | | | | | | | | | | | | | | | | |
| Non-gun owner | 675 | 25.1 [22.6-27.8] | 1249 | 51.2 [48.1-54.3] | 389 | 21.9 [19.3-24.8] | 690 | 26.2 [23.7-29.0] | 1162 | 47.4 [44.3-50.4] | 460 | 24.6 [21.9-27.4] | 2,341 | 100 | 1.1 | -3.8[b] | 2.6 |
| Gun owner, no purchase | 209 | 37.5 [31.1-44.3] | 279 | 55.1 [48.3-61.8] | 29 | 6.9 [4.4-10.8] | 189 | 28.2 [22.9-34.0] | 268 | 54.1 [47.2-60.8] | 60 | 17.4 [12.2-24.2] | 519 | 100 | -9.3[a] | -1.1 | 10.5[a] |
| Gun owner, purchase | 6 | 79.7 [50.1-93.9] | 3 | 15.4 [4.0-44.0] | 1 | 5 [0.6-30.2] | 4 | 68.5 [35.9-89.4] | 4 | 22 [6.2-54.8] | 2 | 9.5 [2.0-34.4] | 10 | 100 | -11.1 | 6.6 | 4.5 |

PD = Prevalence difference; CI = Confidence interval

Rows and columns may not sum to total because of missing values.

Counts are unweighted; percentages and 95% CIs are survey-weighted.

[a]P<0.05

[b]P<0.10
